# Nigrosome-1 neuromelanin-iron coupling profiles in idiopathic and *GBA*-associated Parkinson’s disease

**DOI:** 10.1101/2025.11.03.25339328

**Authors:** Martín Martínez, Mikel Ariz, Ignacio Alvarez, Tomás Muñoz Santoro, Gabriel Castellanos, the Catalonian Neuroimaging Parkinson’s disease Consortium, Katrin Beyer, Yaroslau Compta, Àngels Bayés, Dolores Vilas, Juan Marin, Jorge Hernández-Vara, Carlos Ortiz de Solórzano, Maria A. Pastor, Pau Pastor

## Abstract

**Background:** Firstly, to determine whether iron-sensitive MRI measures mediate neuromelanin (NM) loss within the substantia nigra pars compacta (SNc) and its subregion, nigrosome-1 (N1) in idiopathic Parkinson’s disease (iPD) and *GBA*-associated Parkinson’s disease (GBA-PD) versus healthy controls (HC). Secondly, to assess the diagnostic value of NM-and iron MRI metrics, including their lateralisation.

**Methods:** Eighty-six participants (HC=30, iPD = 30, GBA-PD = 26) underwent midbrain quantitative NM-MRI and susceptibility-weighted imaging (SWI). Contrast ratio (CR), normalised volume (nVol) and left-right asymmetry indices (AI) of SNc and N1 calculated. Receiver-operating analyses assessed group discrimination and onset-side prediction. Bidirectional causal mediation (5,000 bootstraps, FDR-corrected) tested Iron→NM→group and NM→Iron→group pathways. Moderation models examined the effect of GBA-mutation severity.

**Results:** iPD and GBA-PD showed marked NM loss and increased iron in SNc/N1, being more prominent in GBA-PD. N1 NM metrics provided the strongest discrimination (HC vs GBA-PD AUC=0.93, HC vs iPD AUC=0.83), followed by iron measures (AUC=0.78-0.89). N1 NM asymmetry yielded moderate lateralisation accuracy (AUC=0.76). Cross-sectional mediation analyses identified significant Iron→NM→group effects across SNc and N1 (q<0.01), supporting mechanistic hypotheses of iron-driven NM depletion in PD. In GBA-PD, NM-iron coupling in N1 was influenced by mutation severity, suggesting a genotype-specific disruption of NM-iron homeostasis.

**Conclusions:** Multi-contrast MRI centered on N1 reveals an iron-driven NM imbalance that distinguishes PD from controls and it is modulated by *GBA* mutation severity, supporting the applicability of N1-focused NM-iron imaging as a biomarker for PD diagnosis.

Note: This manuscript has been submitted to Journal of Neurology, Neurosurgery & Psychiatry for consideration.

## Introduction

Parkinson’s disease (PD) is a heterogeneous neurodegenerative disorder characterised by motor and non-motor symptoms. The clinical variability across PD patients underscores the need for reliable biomarkers to capture disease mechanisms, improve diagnostic accuracy, and facilitate stratification for targeted interventions.

The progressive loss of dopaminergic neurons containing neuromelanin (NM) within the substantia nigra pars compacta (SNc) is a hallmark of PD [1]. NM plays a central role in long-term iron storage and redox regulation [2]. Motor symptoms emerge after 50-70% dopaminergic cell loss in the SNc, with NM depletion progressing rapidly in early disease stages [3]. Nigrosome-1 (N1), the most vulnerable SNc subregion, has been proposed as a particularly sensitive biomarker of early PD [4,5].

Genetic factors contribute substantially to PD heterogeneity. Variants in the acid β-glucocerebrosidase (*GBA*) gene (MIM:606463) are associated with PD in 10-20% [6]. *GBA* mutation carriers frequently exhibit earlier onset and faster motor progression, compared with idiopathic PD (iPD). These facts suggest that a different vulnerability of dopaminergic and non-dopaminergic neurons in *GBA*-associated PD (GBA-PD), could be reflected in specific NM-and iron-related MRI signatures.

Recent studies have shown that NM-and iron-sensitive MRI analysis can differentiate iPD from healthy controls (HC) [4], and it can indeed distinguish genetic PD subtypes [7]. NM and iron are tightly intertwined under physiological conditions since NM binds and buffers iron, when dysregulated, NM may promote oxidative stress and neuronal injury, supporting underlying bidirectional interactions [2]. PD motor signs start and remain asymmetric as the disease progresses [8,9]. Neuropathology and PET-SPECT studies show greater loss of nigrostriatal neurons and striatal denervation contralateral to the most affected side [10]. A multimodal MRI study in early PD N1 reflects asymmetric N1 NM loss with concomitant iron accumulation contralateral to the most affected side [5].

The present study aimed to address three main questions. Firstly, whether group differences in NM-and iron-sensitive MRI metrics could be detected across the SNc and N1. Secondly, whether asymmetry indices of SNc and N1 NM-iron MRI differentiate iPD from GBA-PD. Thirdly, whether NM and iron metrics are mechanistically coupled through bidirectional mediation pathways, and whether these relationships are further moderated by mutation *GBA* severity.

## Methods

### Study design and participants

A cross-sectional observational study including iPD, GBA-PD and healthy controls (HC) was conducted. Groups were matched for age and sex. All participants underwent standardised neurological assessment, including age, sex, education, disease duration, Hoehn & Yahr (HY) stage, Unified Parkinson’s Disease Rating Scale part III (UPDRS-III), levodopa equivalent daily dose (LEDD), Mini-Mental State Examination (MMSE), handedness, and the hemibody of motor onset. Presence of REM sleep behaviour disorder (RBD) was assessed with the Single-Question Screen for RBD [11]. In GBA-PD, mutation severity was classified as risk, mild, or severe according to the proposed classification [6].

### MRI acquisition and processing

All subjects underwent NM-sensitive MRI and susceptibility-weighted imaging (SWI) as a proxy for iron-sensitive contrast. Regions of interest (ROIs) including the SNc and N1 were automatically segmented using a multi-atlas approach [4,12], (**Supplementary Material)**. For each ROI, contrast ratio (CR) and normalised volume (nVol: raw volume divided by total intracranial volume) were extracted. Bilateral measures were harmonised, and individual-level quality control was performed to ensure reproducibility.

### Outcomes

The primary endpoint was the between-group difference in SNc NM nVol. Secondary outcomes included NM CR and SWI-derived iron-sensitive metrics across all ROIs, as well as asymmetry indices (AIs) derived from bilateral measures.

## Statistical analysis

Descriptive statistics (mean, SD, median, quartiles) were computed for each parameter. Global group differences were assessed with Kruskal-Wallis tests, followed by Wilcoxon pairwise contrasts. Effect sizes were reported as Hedges’ g and Cliff’s delta. Discriminative performance was quantified using receiver operating characteristic (ROC) area under the curve (AUC) and 95% confidence intervals (CIs) by DeLong’s method.

Left-right asymmetry indices (AI) were computed as (R - L)/(R + L) and oriented to the clinically affected (onset) side in patients. In between-group comparisons, AI values were corrected for the median orientation bias observed in HC. Sensitivity analyses included centering AI values against HC medians.

Bidirectional mediation models were tested for NM→Iron and Iron→NM pathways in CR and nVol, estimating indirect (average causal mediation effect, ACME), direct (ADE), and total effects with 5,000 bootstrap simulations. Moderation of NM-iron coupling by GBA mutation severity was tested using interaction terms and simple-slope probes. Covariate-adjusted models included age, sex, dexterity, onset side, disease duration, education, HY stage, LEDD, MMSE, RBD, and UPDRS-III. *GBA* mutation severity was treated as a categorical factor (risk, mild, severe)[6] when testing group-wise differences in NM-iron coupling. Multiple testing was corrected with Benjamini-Hochberg false discovery rate (FDR).

All analyses were two-tailed with α=0.05. Group and mediation models were estimated using ordinary least squares (OLS) regression with heteroskedasticity-consistent (HC2) robust covariance estimators. Analyses were performed with R (v4.2.2; R Foundation for Statistical Computing, Vienna, Austria) using standard packages including *tidyverse* for data wrangling, *ggplot2* for visualisation, *pROC* for ROC analyses, *effsize* for non-parametric effect sizes, and *mediation* for mediation modelling [13]. Additional information is available on sample size and power, participants, genetic analysis, MRI acquisition and processing, MRI derived metrics, and data analyses in the **Supplementary Methods**.

## Results

### Demographic and clinical characteristics

Eighty-six participants were included (HC=30, iPD=30, and GBA-PD=26; **Table 1**). Handedness and motor onset side were similarly distributed in PD groups. Demographic and clinical measures including sex, education, disease onset, disease duration, HY stage, LEDD, MMSE, and RBD prevalence did not differ significantly between patient groups. Statistically significant differences were found for UPDRS-III scores, which were higher in GBA-PD compared with iPD, consistently with the more pronounced motor impairment among GBA-PD (F=5.62, *p*=0.022; **Table 1**).

**Table 1.** Demographic and clinical data of the cohort. Kruskal-Wallis and Chi-square tests were applied to compare categorical variables, whereas heteroscedastic one-way ANOVAs (Welch) were used for continuous variables. All tests were two-sided. Quantitative data are shown as mean±SD, and categorical data as counts. Abbreviations: F, female; M, male; R, right; L, left; A, ambidextrous; B, bilateral; H&Y, Modified Hoehn & Yahr stage; UPDRS-III, Unified Parkinson’s Disease Rating Scale part III; LEDD, levodopa equivalent daily dose; MMSE, Mini-Mental State Examination; RBD, REM behaviour disorder; NA, not applicable/available; HC, healthy controls; iPD, idiopathic Parkinson’s disease; GBA-PD, GBA-associated Parkinson’s.

|  | HC (n=30) | iPD (n=30) | GBA-PD (n=26) | Comparison |
| --- | --- | --- | --- | --- |
| Age | 62.73±7.83 | 64.33±7.65 | 63.69±9.35 | F=0.32, p=0.728 |
| Sex (Female / Male) | 17 / 13 | 14 / 16 | 12 / 14 | $\chi^2=0.82$, p=0.663 |
| Handedness (L / A / R) | 0 / 2 / 28 | 0 / 1 / 29 | 2 / 2 / 22 | $\chi^2=2.96$, p=0.121 |
| Most affected body side (L / B / R) | NA | 13 / 1 / 16 | 16 / 0 / 10 | $\chi^2=2.42$, p=0.121 |
| Years of education | NA | 10.63±5.16 | 12.04±4.07 | F=1.29, p=0.5261 |
| Age at onset | NA | 56.27±9.98 | 55.5±9.31 | F=0.09, p=0.767 |
| Disease duration | NA | 8.07±4.94 | 8.19±5.66 | F=0.01, p=0.930 |
| H&Y stage | NA | 2.07±0.29 | 2.12±0.326 | $\chi^2=0.66$, p=0.416 |
| UPDRS-III | NA | 21.27±8.12 | 28.00±12.3 | F=5.62, p=0.022 |
| LEDD | NA | 718±375 | 632±386 | F=0.70, p=0.405 |
| MMSE | NA | 26.4±2.80 | 26.0±3.54 | F=0.15, p=0.705 |
| RDB (Yes/No) | NA | 18 / 12 | 15 / 11 | $\chi^2=0.01$, p=1 |

### *GBA* gene variants

The GBA-PD cohort comprised 26 heterozygous carriers of the following *GBA* variants: Val54GlnFs*11 (n=1), Ser52Leu (n=1), E326K (n=5), T369M (n=1), N370S (n=4), Tyr244Cys (Y205C) (n=2), R329C (n=1), R496H (n=1), L444P (n=7), G202R (n=1), D409H (n=1), G195W (n=1). Severity totals were: risk-variant (n=6), mild (n=8), severe (n=10), unknown (n=2). Classification of *GBA* variants followed the GBA1-PD browser [6].

**Supplementary Table 1** shows the demographic and clinical data of the GBA-PD group stratified by mutation severity.

### Group-level differences on MRI parameters

Both iPD and GBA-PD groups showed significantly lower NM CR and nVol, as well as higher iron indices in SNc and N1 compared with HC, with the largest differences found at N1 (**Figure 1, Supplementary Table 2**). Global Kruskal-Wallis tests (**Supplementary Table 3**) showed statistically significant differences in NM depletion (CR and nVol) in SNc and N1 of PD groups (e.g., SNc NM nVol: χ²(2)=40.16, q<0.0001; SNc NM CR: χ²(2)=32.14, q<0.0001; N1 NM CR: χ²(2)=34.68, q<0.0001). Iron-sensitive indices were higher in PD, particularly at N1 (N1 Iron CR: χ²(2)=29.47, q<0.0001).

**Figure 1.**
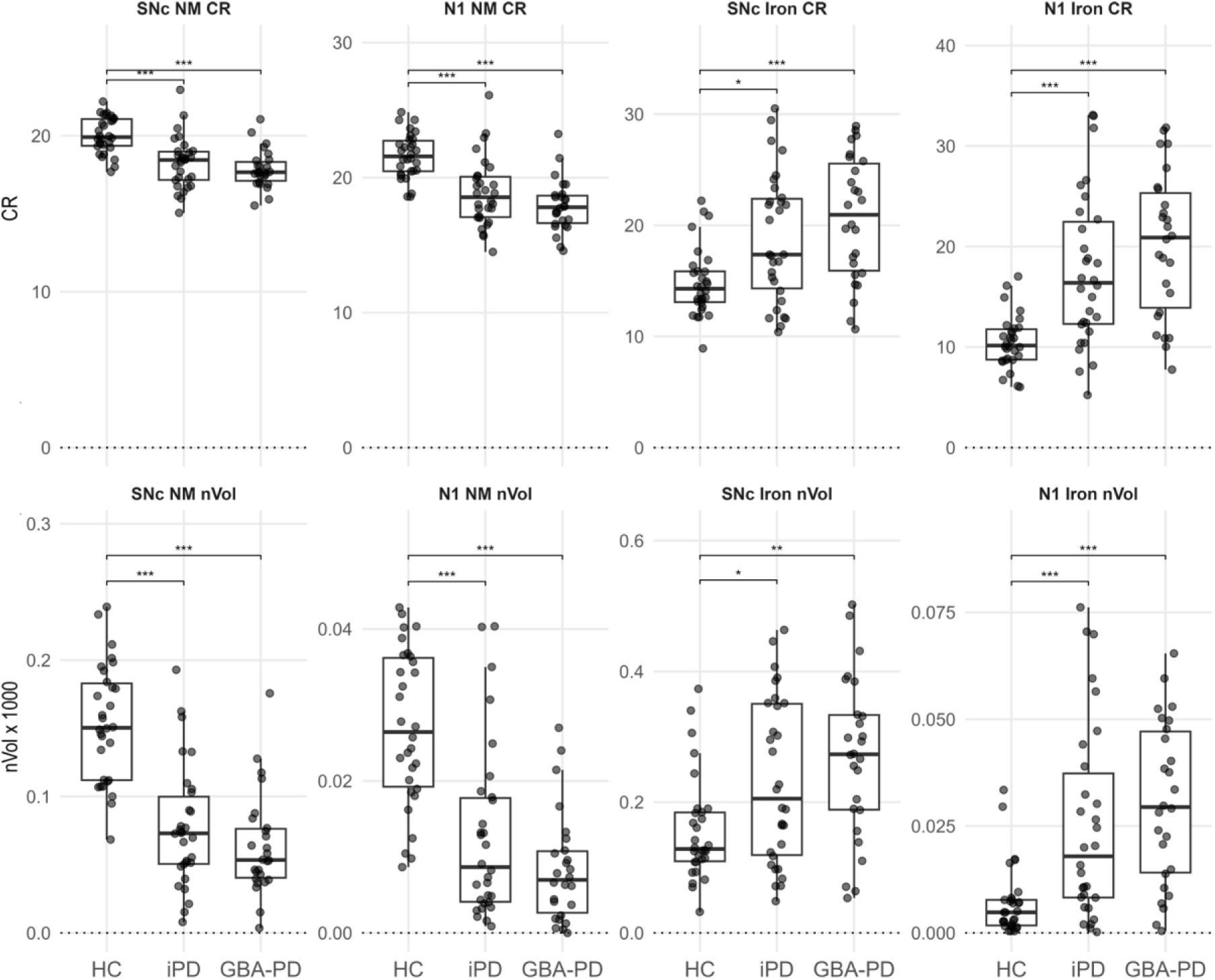
Box plots of the brainstem MRI parameters quantified in the 3D-ABSP. Data is presented with a box plot and individual scatter points of each subject’s quantitative measures (*p < 0.05, **p < 0.01, ***p < 0.001, corrected for multiple testing). Abbreviations: NM, neuromelanin; SNc, substantia nigra; N1, nigrosome-1; nVol, normalised volume; CR, contrast ratio; HC, healthy controls; iPD, idiopathic Parkinson’s disease; GBA-PD, GBA-associated Parkinson’s.

Pairwise post-hoc tests (**Supplementary Table 4**) confirmed that HC showed higher NM CR and nVol than both iPD and GBA-PD in SNc and N1 (e.g., SNc NM CR: HC>iPD, Hedges’ g=1.24, q<0.0001; HC>GBA-PD, g=1.84, q<0.0001; N1 NM CR: HC>iPD, g=1.27, q<0.0001; HC>GBA-PD, g=2.05, q<0.0001). Differences between iPD and GBA-PD were small for NM metrics (e.g., SNc NM CR: g=0.28, q=0.3273). In contrast, iron measures were higher in GBA-PD than HC, being more prominent in N1 (Iron CR: GBA-PD>HC, g=1.82, q<0.0001; Iron nVol: GBA-PD>HC, g=1.61, q<0.0001; negative g indicates higher values in GBA-PD).

Receiver-operating curve values supported these results (**Supplementary Table 4**). Discrimination was strongest for GBA-PD vs HC considering NM-based markers, with high AUC values (SNc NM nVol: 0.933, 95%CI 0.867-1; N1 NM CR: 0.928, 95%CI 0.851-1). NM-derived metrics also provided high discrimination between HC and iPD (SNc NM nVol: 0.891, 95%CI 0.807-0.976; N1 NM CR: 0.832, 95%CI 0.721-0.942). Discrimination between iPD and GBA-PD remained weak (AUC=0.60).

### Asymmetry indices

Since HC showed consistent lateralisation in certain parameters towards the left side (e.g., N1 NM CR and nVol, and SNc and N1 Iron CR), AIs were centered using HC anchors to ensure that the patient-control contrasts would reflect only the pathological asymmetry differences (**Supplementary Table 5**).

Statistically significant group effects across SNc and N1 were confined to N1 NM metrics (**Figure 2**; **Supplementary Table 6**). Global tests were statistically significant for N1 NM CR (χ²(2)=15.6, q=0.0033) and N1 NM nVol (χ²(2)=12.4, q=0.0082), while SNc and iron-based AIs did not reach statistical significance (all q≥0.58). Post-hoc tests showed that PD had a stronger negative AI shift than HC (e.g., N1 NM CR: HC>iPD g=0.95, q=0.008; HC>GBA-PD g=0.84, q=0.008; N1 NM nVol: HC>iPD g=1.01, q=0.008; HC>GBA-PD g=0.87, q=0.044; **Supplementary Table 7**) reflecting a more impaired left brain side despite of the PD onset side. No statistically significant differences emerged between iPD and GBA-PD.

**Figure 2.**
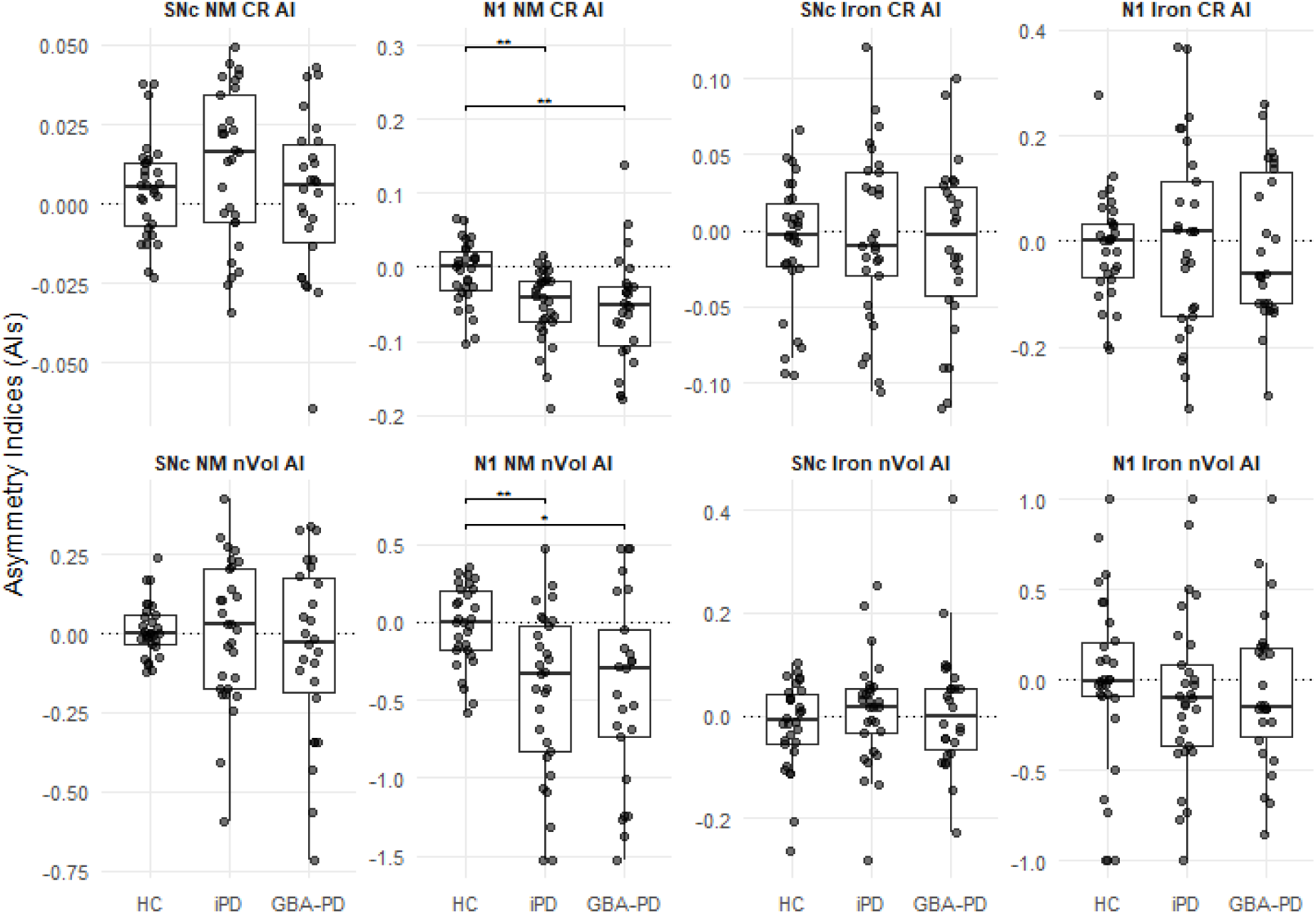
Group differences in asymmetry indices (AI) for SNc and N1. Asymmetry indices (AI) across groups. For each parameter, whether the HC AI distribution differed from zero was tested; when statistically significant, the HC median from all groups (HC-anchored correction) were subtracted. In this dataset the HC-anchor was applied to: N1 NM CR, N1 Iron CR, SNc NM nVol, and N1 NM nVol (see **Supplementary Table 5**). Panels display parameter (CR/nVol) and region×sequence by column (SNc/N1 × NM/Iron). Boxes show medians and IQR; points are individual subjects. Significance brackets reflect FDR-adjusted pairwise Wilcoxon tests (q<0.05 *; q<0.001 **). Abbreviations: NM, neuromelanin; SNc, substantia nigra; N1, nigrosome-1; nVol, normalised volume; CR, contrast ratio; HC, healthy controls; iPD, idiopathic Parkinson’s disease; GBA-PD, GBA-associated Parkinson’s.

Discrimination analyses mirrored these results. N1 NM AI provided moderate accuracy for HC vs PD patients (e.g., CR AUC=0.76 [0.64-0.88] for iPD, and CR AUC=0.76 [0.62-0.89] for GBA-PD; nVol AUC=0.75 [0.62-0.88]) for iPD, and nVol AUC=0.71 [0.56-0.87]) for GBA-PD) but near-chance classification for iPD vs GBA-PD (AUC=0.51-0.61; **Supplementary Table 7**). Onset-stratified summaries confirmed contralateral dominance: in iPD, left brain-onset patients had more negative AIs than right brain-onset (e.g., N1 NM CR mean-0.025 vs-0.002), similar patterns were observed in GBA-PD (**Supplementary Table 8**). These results reinforce the validity of N1 asymmetry as a biomarker of symptom lateralisation with a special sensitivity of the left brain towards degeneration, opposite to HC which showed a higher left side NM nVol and CR.

### Mediation between neuromelanin and iron

Mediation analyses revealed a predominant “Iron → NM → Group” (reverse) pathway within both SNc and N1 in PD vs HC (**Figure 3**; **Supplementary Table 9**). In these models, the indirect effect (ACME) represents the portion of group differences in NM explained by iron-sensitive variance, whereas the direct effect (ADE) reflects residual NM differences not accounted for by iron. A statistically significant ACME thus indicates that iron imbalance partially explains NM loss across groups, while a negative statistically significant ADE suggests additional NM-linked alterations independent of iron.

**Figure 3.**
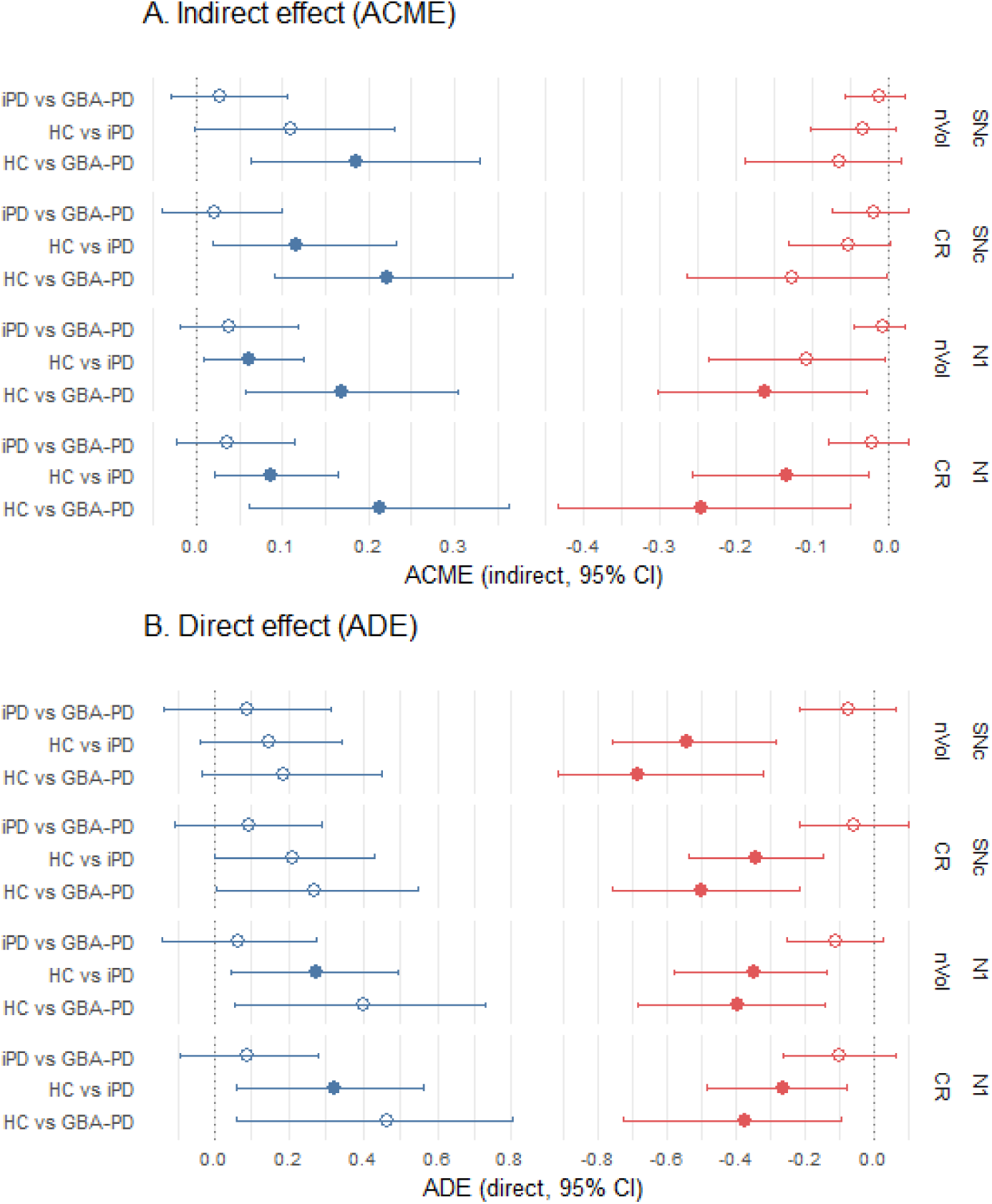
Bidirectional mediation between NM and iron across groups. **A.** Indirect effects (ACME). **B.** Direct effects (ADE). Rows show region × measure (SNc/N1 by nVol, or CR). Columns within each panel split into two causal directions: **Iron→NM→grp (blue)** and **NM→Iron→grp (red)**. The y-axis lists pairwise contrasts (iPD vs GBA-PD, HC vs iPD, HC vs GBA-PD). Points are effect estimates with 95% CIs; open markers denote non-statistically significant. X-axes are displayed on a common scale within each panel (effect size units), centred at 0. Effects were estimated with the mediation package using linear models for mediators and logistic models for the group outcome, with age and sex as covariates. For each contrast, the treatment was defined as a shift from the 25th to the 75th percentile of the putative cause (NM or Iron); 5,000 simulations with bootstrap were attempted (falling back to quasi-Bayesian when needed). Multiple-comparison-adjusted q values (Benjamini-Hochberg) are provided in **Supplementary Table 9**. Abbreviations: ACME, average causal mediation effect; ADE, average direct effect; NM, neuromelanin; Iron (SWI); SNc, substantia nigra; N1, nigrosome-1; nVol, normalised volume; CR, contrast ratio; HC, healthy controls; iPD, idiopathic Parkinson’s disease; GBA-PD, GBA-associated Parkinson’s.

In the HC vs GBA-PD contrast, this pathway showed consistent and FDR-significant indirect effects across regions (e.g., SNc CR ACME = 0.222, q = 0.0016; N1 CR ACME = 0.214, q = 0.006; SNc nVol ACME = 0.186, q = 0.0048; N1 nVol ACME = 0.170, q = 0.0048), with substantial mediated proportions (34-50%). The “NM → Iron → Group” forward direction did not reach FDR significance (q = 0.05-0.11), although direct effects remained negative and statistically significant (SNc CR ADE =-0.500, q = 0.0021; N1 CR ADE =-0.372, q = 0.0064; N1 nVol ADE =-0.393, q = 0.0021). These results suggest that iron acts as an upstream driver of NM differences in GBA-PD, but that persistent NM deficits remain even after accounting for iron, consistent with additional mechanisms such as more intense dopaminergic loss and/or lysosomal dysfunction in GBA-PD.

In the HC vs iPD contrast, smaller but statistically significant mediation appeared selectively in N1 in the “Iron → NM → Group” pathway (ACME = 0.086, q = 0.019 for CR; ACME = 0.061, q = 0.031 for nVol), while direct effects remained significant for the “NM→Iron” models (e.g., SNc CR ADE =-0.343, q = 0.0011; N1 CR ADE =-0.266, q = 0.0052; N1 nVol ADE =-0.349, q = 0.0011). These results suggest that iron contributes to NM loss to a lesser extent, with NM-driven alterations in iPD,.

By contrast, no FDR-significant mediation was found for the iPD vs GBA-PD comparison (all q ≥ 0.22-0.92), indicating that the mechanisms linking NM and iron are qualitatively similar across PD subtypes, though quantitatively more pronounced in GBA carriers.

Collectively, these results indicate a bidirectional but asymmetric NM-iron relationship, with indirect effects predominantly driven by iron-linked variance reflected by NM differences. While the statistical direction suggests that iron-related dysregulation may precede secondary NM loss, the negative ADEs reveal concurrent NM-specific processes, likely reflecting the activation of parallel neurodegenerative cascades not mediated by iron.

NM-iron scatterplots revealed consistent negative coupling across iPD and GBA-PD, in contrast to slightly positive slopes observed in HC (**Figure 4**, Panels A-D, **Supplementary Figure 1 and Supplementary Table 10**). This pattern was evident for both CR and nVol measures, and in both directions (“NM→Iron” and “Iron→NM”), suggesting that PD is characterised by the loss of the physiological covariation between NM and iron burden. The GBA-PD subset exhibited more pronounced slope inversion than iPD, particularly in N1, consistently with an amplified dysregulation of NM-iron homeostasis in genetically driven disease.

**Figure 4.**
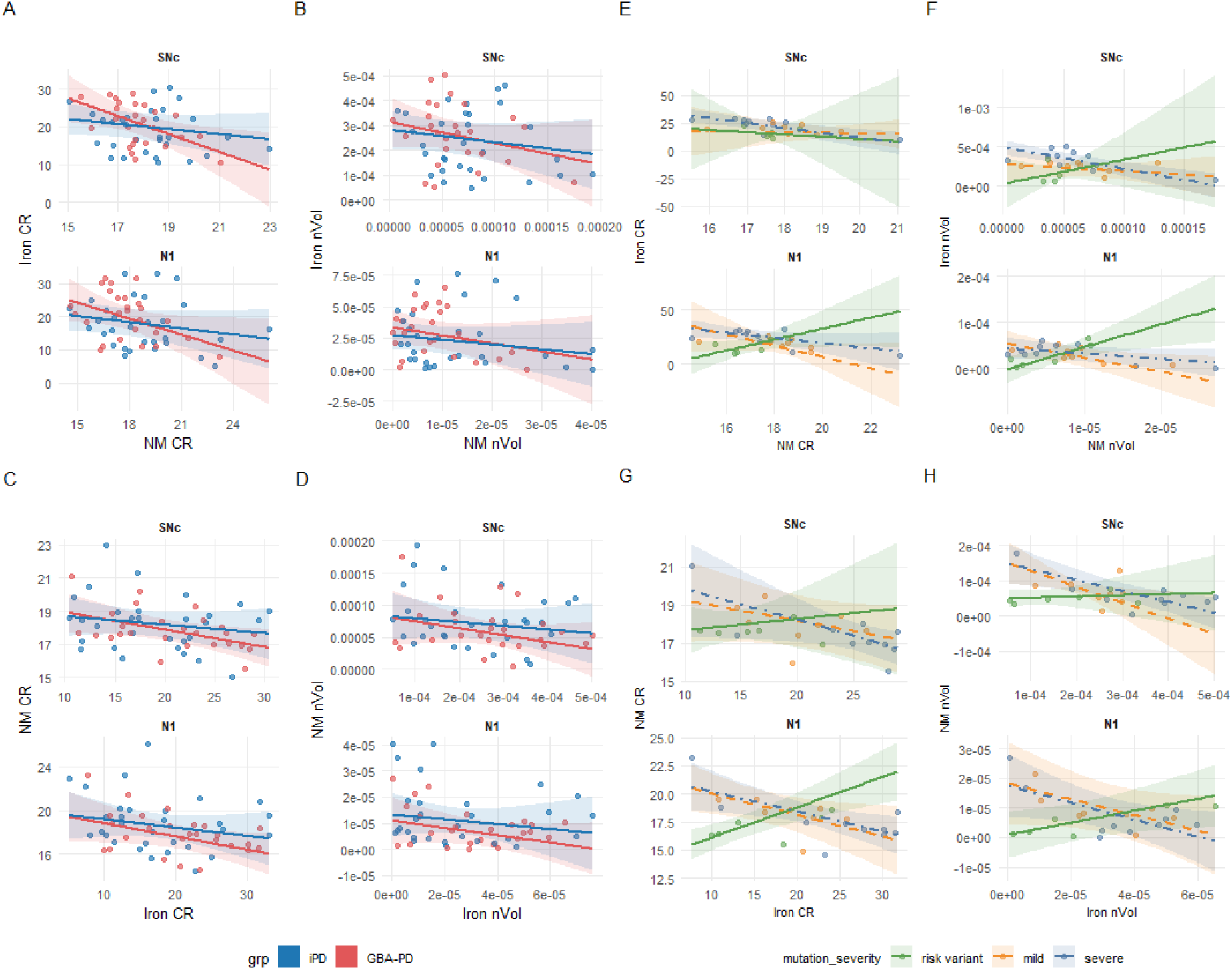
Bidirectional NM-iron coupling in PD and its modulation by GBA mutation severity. Panels A-D illustrate NM-iron coupling across diagnostic groups (iPD, and GBA-PD). Forward direction (A-B) represents NM→Iron coupling; reverse direction (C-D) represents Iron→NM coupling, for both CR and nVol measures. Forward direction (E-F) represents NM→Iron coupling; reverse direction (G-H) represents Iron→NM coupling for both CR and nVol measures in GBA-PD. Lines indicate robust regression fits ± 95 % CIs. Panels E-H show equivalent bidirectional models restricted to GBA-PD patients, stratified by mutation severity (risk-variant, mild, severe). Overall, a negative NM-Iron coupling slope emerges in both PD subgroups, whereas N1 exhibits the strongest and genotype-dependent coupling in GBA-PD, with mild and severe carriers showing attenuated or reversed slopes relative to risk-variant carriers. Abbreviations: NM, neuromelanin; SNc, substantia nigra; N1, nigrosome-1; nVol, normalised volume; CR, contrast ratio; grp, group; iPD, idiopathic Parkinson’s disease; GBA-PD, GBA-associated Parkinson’s. **Abbreviations:** 3D-ABSP = 3D automated MRI atlas-based segmentation pipeline; CR = contrast ratio; *GBA= Acid beta-glucocerebrosidase gene*; GBA-PD = GBA Parkinson’s disease; HC = healthy controls; iPD = idiopathic Parkinson’s disease; LC = locus coeruleus; LEDD = levodopa equivalent daily dose; MRI = Magnetic Resonance Imaging; N1: nigrosome-1; NM = neuromelanin; nVol = normalized volume; PD = Parkinson’s disease; RBD = REM sleep behaviour disorder; RN = red nucleus; SNc = *substantia nigra pars compacta*; UPDRS = Unified Parkinson’s disease Rating Scale.

### Moderation by *GBA* mutation severity

Moderation analyses showed that *GBA* mutation severity selectively modulated coupling at N1 but not at the entire SNc (**Figure 4, Panels E-H**). The forward “NM→Iron” direction revealed a statistically significant interaction for N1 nVol [F(2,9)=5.89, p=0.023, partial R²=0.57] and a trend for N1 CR [F(2,9)=2.99, p=0.10, partial R²=0.40]. Post-hoc slopes indicated preserved positive coupling in risk variant *GBA* carriers (β=4.93 ± 4.35, p=0.29) but attenuated or reversed relationships in carriers of mild (β =-3.09 ± 2.18, p=0.19) and severe *GBA* variants (β =-1.17 ± 1.69, p=0.51). The reverse (Iron→NM) direction showed even stronger moderation in N1 CR (F(2,9)=11.41, p=0.003, partial R²=0.72) and significant effects in N1 nVol (F(2,9)=5.69, p=0.025, partial R²=0.56), reflecting positive slopes in risk-variant *GBA* carriers and negative in mild/severe variant *GBA* carriers. SNc-wide analyses followed a similar pattern but did not reach significance (all p > 0.18).

Together, these results delineate a graded loss of NM-iron coupling strength across diagnostic and genetic dimensions: (1) HC = weak positive coupling; (2) iPD = moderate inversion; (3) GBA-PD = strong inversion modulated by mutation severity. This continuum supports a transition from an NM-supported to an iron-driven regime, where an increase of lysosomal dysfunction could lead to disrupt metal sequestration and NM depletion.

In HC-only models, no statistically significant mediation effects were observed (all q > 0.05), confirming that NM-iron coupling was specific to PD (**Supplementary Figure 1; Table 11**). Sensitivity analyses, including minimal, full, and unadjusted covariate sets, reproduced the main mediation results with less than 10% variation in indirect slopes and consistent ACME/ADE values (**Supplementary Table 12**). Sensitivity analyses confirmed that mediation and moderation results remained consistent after covariate adjustment, alternative scaling of iron-sensitive measures (CR and nVol), and heteroscedasticity-robust estimation (HC2).

## Discussion

This study provides convergent evidence that NM and iron-sensitive MRI sequences of the SNc, particularly in N1, capture disease-relevant alterations in PD, with greater amplification in patients carrying *GBA* mutations. The results of combining group contrasts, asymmetry indices, slope analyses, and bidirectional mediation/moderation models, suggest a mechanistic and clinically meaningful view of NM-iron interplay with mediation pointing predominantly to an Iron→NM→group pathway, especially among GBA-PD.

Both iPD and GBA-PD showed robust NM loss and increased iron-sensitive indices, with the largest effects found in N1, consistent with neuropathological studies identifying this subregion as a primary locus of dopaminergic vulnerability in PD [3,14,15]. The magnitude of iron-related alterations was generally greater in GBA-PD, consistent with the more severe *GBA* phenotypes [16,17]. Direct NM and iron comparisons between iPD and GBA-PD yielded small effects likely reflecting inter-individual heterogeneity and partial overlap of MRI profiles across both PD subtypes. By contrast, group effects were robust for volume-based indices, supporting the consistency of these alterations in GBA-associated PD.

We further investigated whether asymmetry was involved in the PD neurodegeneration. N1 NM asymmetry emerged as a reliable discriminator of patients vs controls, whereas SNc-wide and iron-based asymmetry did not reach significance. Analyses which accounted for the body side where the motor signs start confirmed contralateral dominance of imaging abnormalities, indicating that AIs reflect the clinical lateralization of PD [9,18,19]. Since healthy controls also exhibited small lateralisation biases for a subset of indices, patient AIs were anchored by subtracting the HC distribution deviated from zero, which preserved the qualitative patient-control pattern and enhanced pathological asymmetry [8].

N1 NM asymmetry provided moderate discrimination between patients and controls, but did not differentiate PD subgroups, consistent with convergent nigral vulnerability across PD etiologies [15]. This observation extends prior MRI evidence identifying the “swallow-tail” appearance of N1 as a sensitive imaging marker of nigral integrity in PD [20], suggesting that the present quantitative NM-based asymmetry reflects asymmetric dopaminergic depletion rather than morphological loss. Thus, N1 NM asymmetry primarily indexes shared pathophysiological processes underlying both idiopathic and genetic PD.

Our mediation framework models examined both causal directions (Iron→NM→group and NM→Iron→group). Consistently, FDR-significant ACME effects emerged in the Iron→NM→group path, most prominently in HC vs GBA-PD across SNc and N1 and for both CR and nVol (mediated proportions 30-50%). By contrast, the NM→Iron→group direction did not yield FDR-significant ACME; instead, we observed robust negative ADE in several panels, indicating residual NM-linked group differences not mediated uniquely by iron. This cross-sectional mediation pattern aligns with longitudinal evidence indicating that nigral iron accumulation precedes NM loss in early disease stages, which is consistent with a directional Iron→NM relationship [21,22], and with recent biophysical and quantitative MRI models describing NM-metal interactions and their impact on MR contrast [23,24]. Together, these findings support an upstream role of iron variance in shaping NM loss and group differences.

To better interpret these mediation effects, we examined age-and sex-adjusted HC2-robust slopes of NM-iron coupling across groups. HC showed a positive coupling trend for the Iron→NM direction in N1 nVol, even though it was not statistically-significant after FDR (Supplementary Table 11), patients’ analysis inverted this relationship. The strongest negative slope appeared in GBA-PD for NM→Iron in SNc CR and similar tendency in N1 CR, consistent with a dysregulated NM-iron homeostasis [1,2,25,26].

These slope patterns provide a microstructural substrate for the mediation findings: where group contrasts are largest and coupling is most inverted, Iron→NM yields the strongest indirect effects. This pattern aligns with longitudinal evidence showing that iron accumulation precedes NM loss in early disease stages, suggesting a sequential iron-driven disruption of NM synthesis and dopaminergic integrity [26]. Although NM-iron correlations have been mapped [26] mapped in the anteromedial SNc (N2), quantitative voxel-based morphometry identified colocalized NM loss and iron deposition in posterolateral nigrosomal regions corresponding to N1 and N2 [27]. Together, these observations strengthen the notion that iron dysregulation acts upstream, reshaping NM-iron coupling as reflected in the group-level mediation pattern observed in the present work.

Exploratory analyses indicated that *GBA* gene mutation severity moderated NM-iron coupling selectively within N1. Forward (NM→Iron) N1 nVol showed a significant interaction and N1 CR a strong trend, with differential effects according to the severity of the GBA gene variant: risk-variant carriers retained positive slopes, whereas mild and severe carriers exhibited attenuated or reversed relationships. The reverse (Iron→NM) effect was strongest in N1 CR and also present in N1 nVol, while whole SNc panels were not significant. Although exploratory, these moderation effects were methodologically robust, surviving HC2 correction and showing consistent directionality across NM and iron metrics. The N1-specific pattern aligns with established genotype-phenotype gradients described for GBA-PD [28–30], and suggests that genetic severity may reshape the NM-iron balance beyond absolute signal magnitude.

A previous study using radiomic analyses of NM-and T2*-weighted MRI did not detect significant differences between idiopathic and GBA-associated PD [21], likely because it was focused on static voxel-wise intensity and texture metrics rather than inter-regional relationships. While radiomics captures local heterogeneity, it does not model the functional coupling between NM-and iron-sensitive signals. By contrast, our slope-based approach quantifies their directional interdependence, providing a more dynamic index of metal-handling efficiency. The stronger genotype-related trends found in the present study indicate that the NM-iron interaction itself, rather than its isolated magnitude or texture, may better reflect GBA-driven pathological variability.

Mechanistically, the observed NM-iron coupling pattern is consistent with variant-dependent impairment of lysosomal and metal-handling pathways, since reduced glucocerebrosidase activity compromises lysosomal degradation and lipid recycling, processes tightly coupled to NM synthesis and iron sequestration [25]. When this buffering equilibrium fails, redox-active iron accumulates and oxidative stress rises, leading to the reversal of NM-iron coupling observed in mild and severe variants. The N1 localization of this moderation aligns with its already known dopaminergic vulnerability and with recent high-field and longitudinal MRI studies showing progressive iron loading and NM decline within nigrosome [21,23]. Together, these exploratory findings delineate a continuum in which risk *GBA* variants preserve near-physiological NM-iron coupling, whereas mild and severe *GBA* variants shift toward an iron-driven as well as an NM-depleting regime, reflecting an escalating lysosomal dysfunction and metal dyshomeostasis in GBA-related PD. This reinforces that risk variants are biologically relevant representing an intermediate state within the *GBA* mutation spectrum, consistent with the graded shift in NM-iron coupling observed according to the *GBA* mutation severity [31]. Although the bidirectional mediation models provide mechanistic insights consistent with iron-driven NM depletion, the cross-sectional nature of this study precludes strong causal inference though, owing to the small sample size, these results should therefore be interpreted cautiously and replicated in larger genetic cohorts.

## Data availability

All anonymised data, scripts, and supplementary materials (detailed pipelines, atlas-level results, extended asymmetry tables, mediation outputs, and raw symmetry metrics) will be available in the Supplementary Material or in an open repository (OSF) upon acceptance.

## Ethics approval

Written informed consent was obtained from all participants, and the study protocol was approved by the University of Navarra Research Ethics Committee (Ref. 126/2019).

## Supplementary Material

### Supplementary Methods Sample size and power

The primary endpoint was the between-group difference in SNc NM nVol. Using the smallest reported effect size from prior work (Cohen’s d = 1.20), α = 0.05 (two-tailed), and power = 0.80, the required sample size was 25 per group, allowing 15% for attrition yielded a target of 29 participants per group. Recruitment was balanced across HC, iPD, and GBA-PD.

### Participants

All individuals with PD fulfilled the *UK Brain Bank Parkinson’s disease* criteria [1]. HC without any neurological condition were recruited among PD spouses’.

Patients were invited to attend a visit including an interview recording disease information, demographic data, family history of neurological diseases, and neurological exam. PD patients also underwent a Mini-Mental State Examination (MMSE) [2], the Unified Parkinson’s Disease Rating Scale (UPDRS-III) [3], which was assessed in ON-state REF, Modified Hoehn and Yahr Scale (HY) [4], and Edinburgh Handedness Inventory [5]. Levodopa equivalent daily dose (LEDD) [6]. Disease onset was considered as the age when the parkinsonian signs started. RBD was recorded with the REM Sleep Behavior Disorder Single-Question Screen [7]. The side of onset was harmonised to left/bilateral/right. The selection of individuals of the iPD and HC groups was carried out in reference to the GBA-PD group and controls in order to match sex and age.

### Genetic analysis

Genomic DNA was isolated from peripheral leukocytes. GBA sequencing was carried out by long-range PCR with confirmatory Sanger sequencing [8]. Mutations were classified using the GBA1-PD browser as risk, mild, or severe; variants with scarce information were labelled unknown according to the GBA browser classificator [9].

### MRI acquisition and processing

All MRI scans were acquired on a 3T Siemens MAGNETOM Skyra (Siemens Healthineers, Erlangen, Germany) using a 32-channel head coil. For neuromelanin-sensitive imaging, a 3D T1-weighted gradient echo sequence with magnetization transfer was employed (TR = 34 ms, TE = 4.91 ms, flip angle = 20°, voxel size = 0.6 × 0.6 × 1.0 mm³, FOV = 220 mm, 40 slices, slice thickness = 1 mm with 0.2 mm gap, acquisition time ≈ 4 min). For iron-sensitive imaging, a susceptibility-weighted (SWI) 3D GRE sequence with filtered phase information was used (TR = 28 ms, TE = 20 ms, flip angle = 15°, voxel size = 0.7 × 0.7 × 2.0 mm³, FOV = 220 mm, 20 slices, slice thickness = 2 mm, acquisition time = 2 min). To improve signal-to-noise ratio and minimise motion effects, three repetitions of each NM and SWI sequence were acquired and averaged offline.

Segmentation of the SNc and N1 was performed using a multi-atlas approach with weighted label fusion, adapted from our previous work [10,11]. Individual images were non-linearly registered to the images forming the atlas, which included manually annotated brainstem structures. Transformed labels were then fused, weighted by the similarity between the target image and each atlas image, to obtain the final segmentation of the structures of interest. Labels for N1 were defined in SWI space by expert raters and co-registered with NM images. For each region of interest, contrast ratio (CR) and normalized volume (nVol) were computed as proxies of neuromelanin and iron content. Dice Similarity Coefficients values of automatic segmentation were 0.915 for brainstem, 0.797 for SNc, and 0.642 for N1.

### MRI-derived metrics

We analysed CR and nVol across SNc and N1. Bilateral measures were harmonised to canonical L/R variables.

### Asymmetry index (AI) and orientation

AI was defined as (R-L)/(R+L), where R is anatomical right and L is anatomical left. Orientation was coded as Left (AI<0) or Right (AI>0). In patients, AI distributions were stratified by clinical onset side (left vs right) without sign-flipping to preserve contralateral dominance. When the HC AI distribution significantly differed from zero, we centered all groups by subtracting the HC median for that parameter. Subject-level L/R and AI orientation values were retained in their native form (without sign-flipping) to ensure methodological transparency.

### Group-level summaries and discrimination tests

Descriptives included mean, SD, median, Q1-Q3. Group comparisons used Kruskal-Wallis tests with Benjamini-Hochberg false discovery rate (BH-FDR). Pairwise Wilcoxon tests reported adjusted p-values and effect sizes (Hedges’ g, Cliff’s delta). ROC curves were computed for discriminative contrasts. When AUC < 0.5, the complementary balanced AUC* = max(AUC, 1-AUC) was also reported for interpretative symmetry between NM and SWI contrasts.

### Mediation analyses

Bidirectional mediation analyses were implemented to model the mutual dependencies between NM and iron-sensitive MRI markers. Rather than assuming a unidirectional effect, we tested both NM→Iron and Iron→NM pathways within the same analytical framework. In each model, the predictor (X) and outcome (Y) were swapped, while the indirect path (M, group) captured the opposite MRI modality. This bidirectional design allows distinguishing whether variations in iron are a downstream effect of NM loss (reflecting impaired storage or buffering) or conversely, whether iron accumulation drives NM depletion through oxidative toxicity. Each mediation model quantified three components: (1) the direct path (c′): the relationship between X and Y controlling for M, (2) the indirect path (a×b): the degree to which M mediates that relationship, and (3) the total effect (c).

This bidirectional framework provides a mechanistic insight into the dynamic balance between NM synthesis/storage and iron dysregulation, enabling inferences about whether PD or GBA-related pathology primarily reflects a failure to sequester iron or a secondary degeneration due to iron-driven oxidative stress. Conceptually, mediation analyses identify which component of the NM-iron system acts as the primary driver. A significant NM→Iron mediation implies that NM neuromelanin loss leads to iron release (a storage failure), while a significant Iron→NM mediation suggests that iron overload contributes to NM depletion (a toxic overload). The coexistence of both effects in SNc or N1 would indicate a bidirectional feedback loop, consistent with a self-amplifying neurodegenerative process in dopaminergic neurons.

Bidirectional mediation models were run for NM and iron in SNc and N1 (CR and nVol). Group was the outcome (binary contrasts HC vs iPD, HC vs GBA-PD, iPD vs GBA-PD). Models estimated average causal mediation effect (ACME), and average direct effect (ADE), total effect, and proportion mediated with 5,000 bootstrap simulations. FDR correction was applied across tests. Parallel analyses included eligible covariates; covariate usage and coefficients were exported.

### Moderation analysis

In the GBA-PD group, the models were further tested for moderation by mutation severity, allowing the slope of NM-iron coupling to vary across “risk-variant”, “mild”, and “severe” carriers. Interaction models included severity × metric terms. The results were reported as interaction coefficients, omnibus p-values, and simple slopes by severity class. All effects were estimated with bootstrapping (5,000 samples) and corrected for multiple comparisons (Benjamini-Hochberg).

### Sensitivity analyses

To assess robustness, all primary mediation and moderation models were repeated with full covariate adjustment (age, sex, and dexterity in analyses with HC, while including onset side, disease duration, education, HY stage, LEDD, MMSE, RBD, and UPDRS-III in analyses with PD). Analyses followed a complete-case approach: variables with ≥80% observed data and non-zero variance were retained in each model.

### Data harmonisation and quality filters

Clinical covariates considered were age, sex, handedness, onset, disease duration, education, UPDRS-III, LEDD, HY, MMSE, and RBD. A 80% non-missing threshold and non-zero variance criterion determined eligibility. Complete-case analyses were applied per test. Random seed = 1234 for reproducibility.

### Formatting and export

All outputs were consolidated into a single Excel workbook with per-cell number formats (scientific vs fixed decimal depending on metric). Mirror CSV files were also exported. Raw and processed data, together with the full R pipeline, will be available at OSF/Zenodo (Supplementary docs) upon acceptance.

## Supplementary Results

### GBA-named according to HGVS or to the ancillary nomenclature

Twenty-six patients were heterozygous carriers for one the following *GBA* mutations (GBA-named according to HGVS or to the ancillary nomenclature): c.160delinsCAGTA (p.Val54GlnFs*11) (n=1), c.155C>T (p.Ser52Leu) (n=1); c.1093G>A (p.Glu365Lys) or E326K (n=5), c.1223C>T - c.721G>A (p.Thr408Met) or T369M (n=1), c.1226A>G (p.Asn409Ser) or N370S (n=4), c.731A>G (p.Tyr244Cys) or Y205C (n=2), c.1102C>T (p.Arg368Cys) or p.R329C (n=1), and c.1604G>A (p.Arg535His) or R496H (n=1), c.1448 (p.Leu483Pro) or L444P (n=7), (p.Gly241Arg) or G202R (n=1), c.1342G>C (p.Asp448His) or D409H (n=1), c.700G>T (p.Gly234Trp) or G195W (n=1). **Supplementary Table 1** shows the demographic and clinical data of the GBA-PD group differentiated by mutation severity, resulting in 6 risk-variant, 8 mild, 10 severe and 2 unknown GBA-PD.

### Group level differences

Full descriptive statistics for all MRI parameters (NM and SWI, CR and nVol, across SNc, and N1) are provided in **Supplementary Table 2**. Kruskal-Wallis tests (**Supplementary Table 3**) confirmed significant global differences for all NM measures in SNc and N1 (all q<0.001). Iron-sensitive metrics were also elevated in patients, with strongest effects in N1 (Iron CR χ²=29.5, df=2, q<0.0001).

Pairwise comparisons (**Supplementary Table 4**) showed that healthy controls (HC) consistently exhibited higher NM signal and volume than both iPD and GBA-PD patients in SNc and N1 (all g≥1.2, q<0.0001). Differences between iPD and GBA-PD were small and not significant across NM metrics. In contrast, iron-sensitive measures were higher in GBA-PD relative to HC, most clearly in N1 Iron CR (g=-1.82, q<0.0001).

ROC analyses are reported in **Supplementary Table 4**. Discrimination was strongest for GBA-PD vs HC using N1 Iron CR (AUC=0.887, 95%CI 0.795-0.979) and N1 Iron nVol (AUC=0.863, 95%CI 0.761-0.965). NM-based metrics also discriminated well after direction balancing (e.g., SNc NM nVol balanced AUC=0.93). In contrast, discrimination between iPD and GBA-PD was weak (AUC range 0.54-0.61).

Visual summaries (**Fig. 1**) corroborated these findings: HC clustered highest on NM CR/nVol across SNc/N1, while patients showed higher SWI values in SNc/N1.

### Asymmetry indices

Descriptive statistics for all ROIs and modalities are provided in **Supplementary Table 5**. Global Kruskal-Wallis results are detailed in **Supplementary Table 6**, and pairwise comparisons with effect sizes and FDR corrections and ROC analyses in **Supplementary Table 7**].

Orientation summaries by disease onset (**Supplementary Table 8**) showed that patients displayed contralateral asymmetry aligned with onset side, while controls demonstrated small systematic biases that were corrected through HC anchoring (**Supplementary Table 5**).

### Mediation and moderation analyses

Complete mediation analyses corroborated that NM-iron coupling is most evident when comparing patients with controls, with larger effects in GBA-PD. In unadjusted models (**Supplementary Table 9**), the Iron→NM→group pathway showed FDR-significant indirect effects across SNc and N1 in the HC vs GBA-PD contrast (e.g., SNc CR ACME=0.22, q=0.002; N1 CR ACME=0.21, q=0.006; SNc nVol ACME=0.19, q=0.005; N1 nVol ACME=0.17, q=0.005), with mediated proportions in the 30-50% range. The NM→Iron→group direction concurrently retained significant direct effects (ADE) in SNc and N1, indicating NM-driven differences not fully explained by iron. In HC vs iPD, mediation was smaller but detectable in N1 (CR and nVol; ACME q<0.05), while iPD vs GBA-PD showed no FDR-significant mediation in either direction (all q≥0.160).

Severity moderation analyses within GBA-PD demonstrated region-specific modulation in N1. Forward models (NM→Iron) revealed a significant interaction for N1 nVol (F(2,9)=5.89, p=0.023, partial R²=0.567) and a trend for N1 CR (F(2,9)=2.99, p=0.101, partial R²=0.399), with positive slopes in risk carriers and negative slopes in mild/severe. Reverse models (Iron→NM) confirmed robust moderation in N1 CR (F(2,9)=11.4, p=0.003, partial R²=0.717) and N1 nVol (F(2,9)=5.69, p=0.025, partial R²=0.558). Pairwise Δslopes reached nominal significance for risk vs mild and risk vs severe in N1 panels, with consistent directionality across forward and reverse specifications (**Supplementary Table 10**). By contrast, SNc panels did not show significant moderation by severity.

### Sensitivity analyses

To evaluate model robustness, all mediation analyses were repeated with unadjusted, minimal, and full covariate sets. No significant moderation effects in NM-coupling were detected in the HC-only group (all q > 0.05), as shown in **Supplementary Table 11**, supporting the specificity of NM-iron coupling to Parkinsonian cohorts. Estimates of indirect (ACME), direct (ADE), and total effects remained consistent across specifications (**Supplementary Table 12**).

## Data Availability

All data produced in the present study are available upon reasonable request to the authors
All data produced in the present work are contained in the manuscript
All data produced are available online at request

---

**Supplementary Figure 1.**
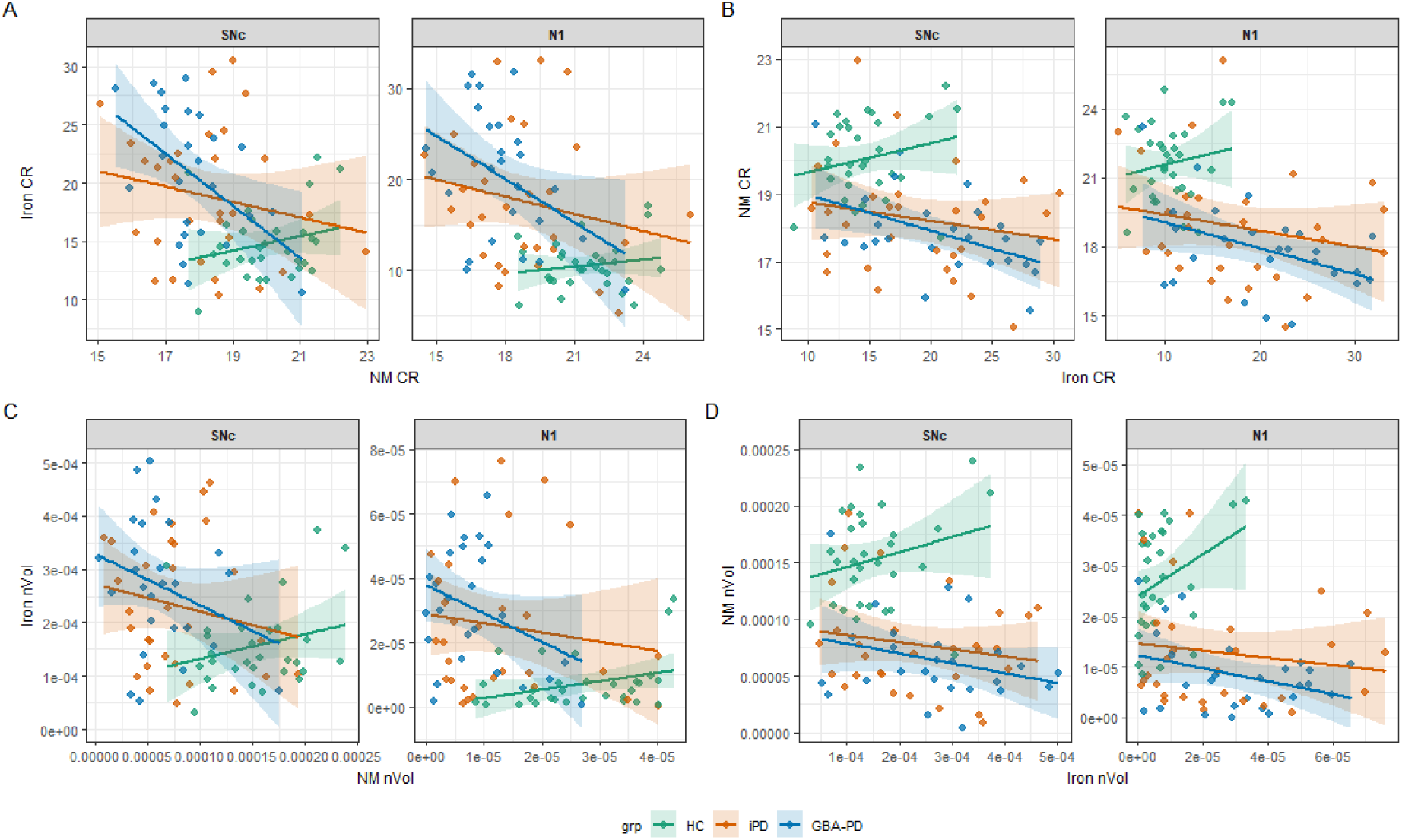
Neuromelanin-iron coupling across HC, iPD, and GBA-PD groups (robust OLS + HC2). Scatterplots showing the relationship between NM-and iron-sensitive MRI indices in the SNc and N1. Panels A-C display forward coupling (NM → Iron) using CR and nVol indices. Panels B-D depict reverse coupling (Iron → NM) using CR and nVol indices. Each subplot shows robust ordinary least squares (OLS) fits with heteroskedasticity-consistent (HC2) standard errors. Green, orange, and blue denote healthy controls (HC), idiopathic Parkinson’s disease (iPD), and GBA-associated PD (GBA-PD), respectively. Shaded ribbons represent 95 % confidence intervals. Rows correspond to imaging measures (top: CR; bottom: nVol) and columns to coupling direction (left: NM → Iron; right: Iron → NM). This figure illustrates the full-group NM-iron coupling profiles complementing the pairwise contrasts reported in Supplementary Table 11. Abbreviations: HC2, heteroscedasticity-robust estimation; NM, neuromelanin; SNc, substantia nigra; N1, nigrosome-1; nVol, normalised volume; CR, contrast ratio; grp, group; HC, healthy controls; iPD, idiopathic Parkinson’s disease; GBA-PD, *GBA*-associated Parkinson’s.

**Supplementary Table 1.** Demographic and clinical data of the GBA-PD differentiated by mutation severity. Kruskal-Wallis and Chi-square tests were applied to categorical variables; heteroscedastic one-way ANOVAs (Welch) were used for continuous variables. All tests are two-sided. Quantitative data are mean±SD; categorical data are counts. Abbreviations: GBA-PD, GBA-associated Parkinson’s; F, female; M, male; R, right; L, left; A, ambidextrous; B, bilateral; H&Y, Modified Hoehn & Yahr stage; UPDRS-III, Unified Parkinson’s Disease Rating Scale part III; LEDD, levodopa equivalent daily dose; MMSE, Mini-Mental State Examination; RBD, REM behaviour disorder; NA, not applicable/available. Unknown severity is excluded from the statistical comparison.

| **Variable/Mutation Severity** | **Unknown (n=2)** | **Risk variant (n=6)** | **Mild (n=8)** | **Severe (n=10)** | **Group comparison\*** |
| --- | --- | --- | --- | --- | --- |
| Age | 60±18.4 | 66.5±7.26 | 62.6±8.16 | 63.6±10.7 | $F=0.44, p=0.651$ |
| Sex (Female / Male) | 1 / 1 | 3 / 3 | 4 / 4 | 4 / 6 | $\chi^2=0.23, p=1$ |
| Handedness (L / A / R) | 0 / 0 / 2 | 0 / 0 / 6 | 2 / 0 / 6 | 0 / 0 / 10 | $\chi^2=4.11, p=0.165$ |
| Most affected body size (L / B / R) | 1 / 0 / 1 | 2 / 0 / 4 | 5 / 0 / 3 | 8 / 0 / 2 | $\chi^2=3.48, p=0.189$ |
| Years of education | 13.5±0.71 | 9.67±3.39 | 14.8±4.10 | 11±3.83 | $F=3.29, p=0.070$ |
| Age at onset | 58±18.4 | 59.8±9.15 | 51±7.73 | 56±8.84 | $F=1.87, p=0.196$ |
| Disease duration | 2±0 | 6.67±3.93 | 11.6±6.76 | 7.6±4.88 | $F=1.45, p=0.270$ |
| H&Y stage | 2±0 | 2±0.55 | 2.12±0.23 | 2.2±0.258 | $\chi^2=0.59, p=0.745$ |
| UPDRS-III | 11.5±2.12 | 25.7±9.77 | 28.6±4.66 | 32.2±16.4 | $F=0.48, p=0.632$ |
| LEDD | 325±106 | 614±332 | 632±351 | 705±477 | $F=0.11, p=0.901$ |
| MMSE | 24±4.24 | 27.3±2.07 | 26.9±3.87 | 25±3.86 | $F=1.20, p=0.332$ |
| RDB (Yes/No) | 1 / 1 | 3 / 3 | 5 / 3 | 6 / 4 | $\chi^2=0.24, p=1$ |

**Supplementary Table 2.**
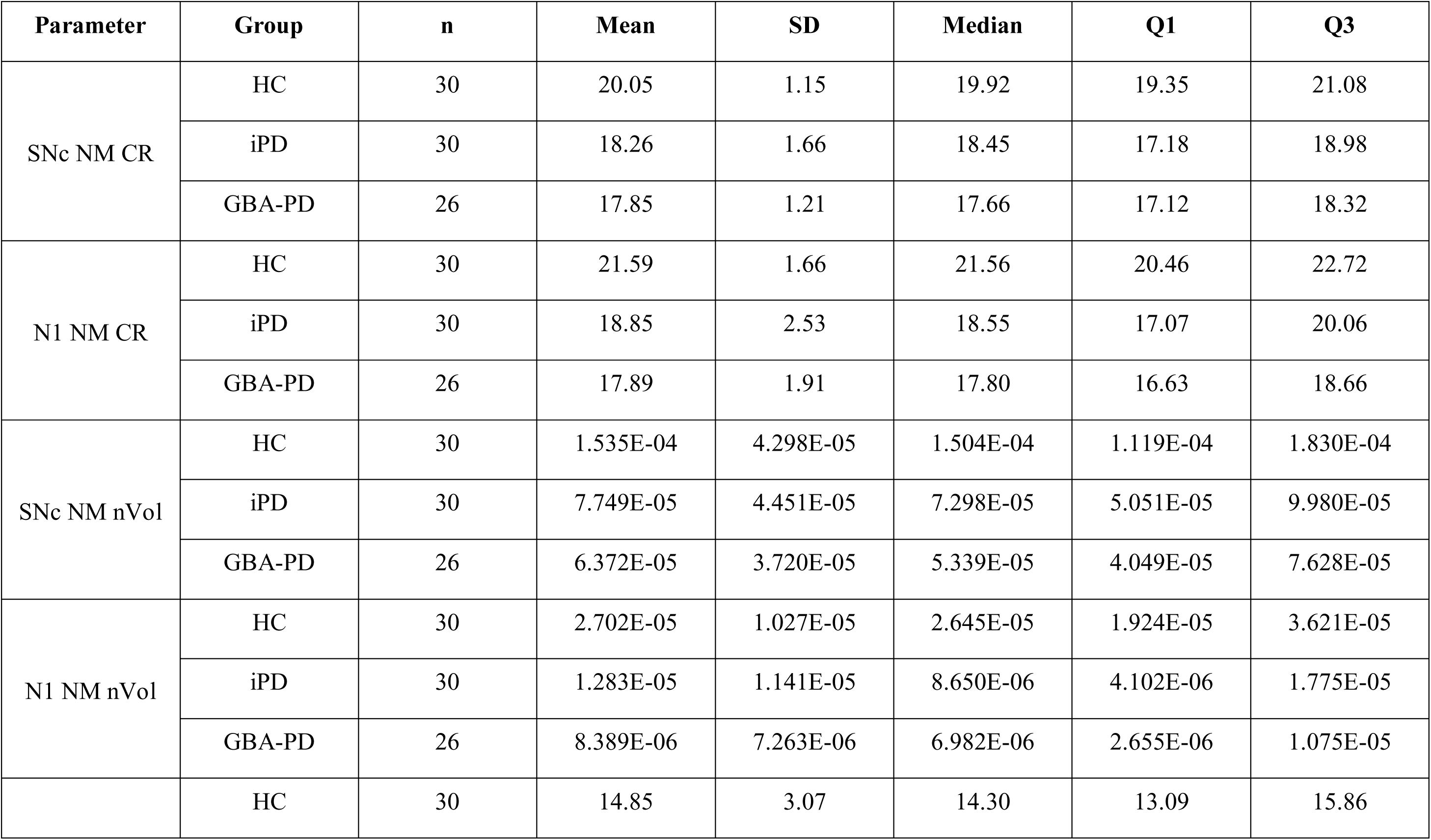
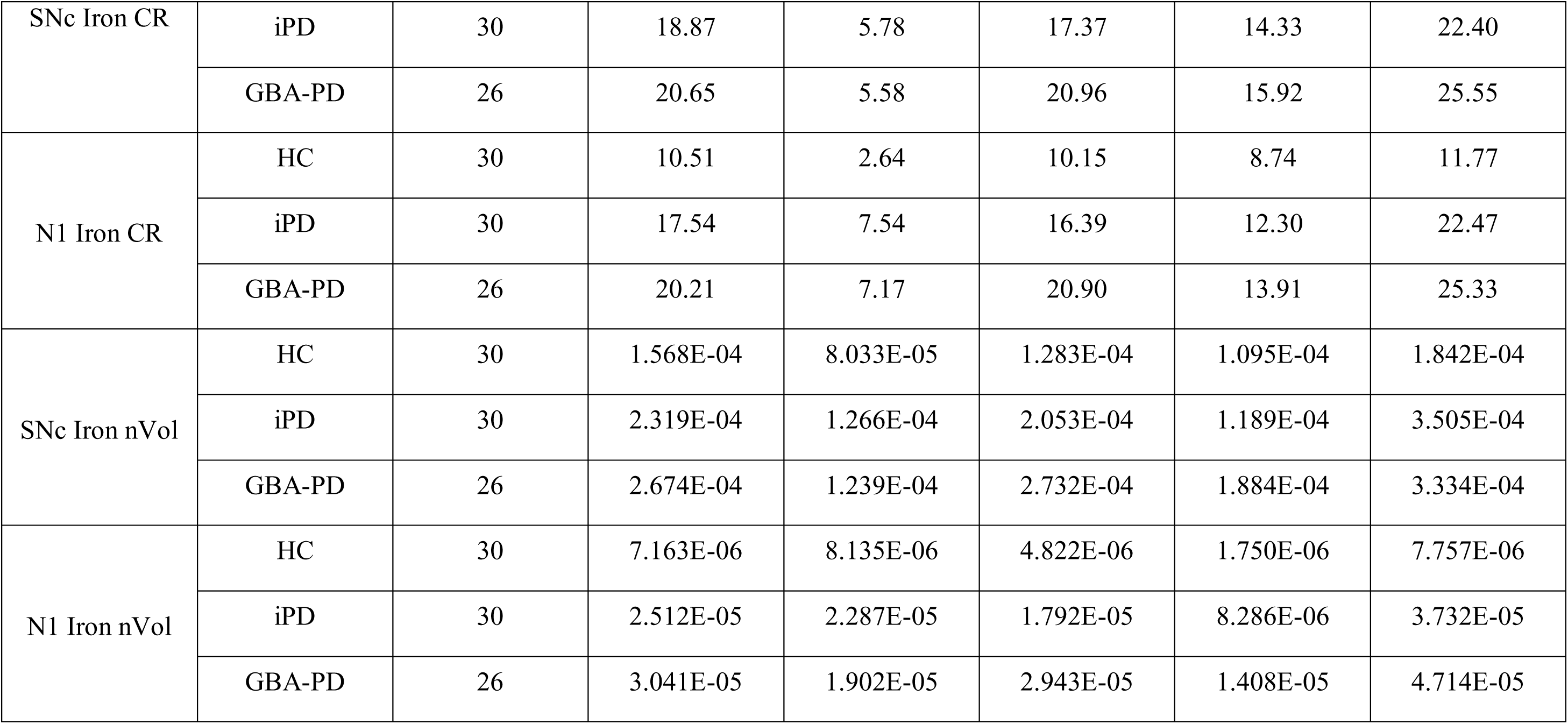
Descriptive statistics of NM-and iron-sensitive MRI parameters across groups. Values represent mean, standard deviation (SD), median, first quartile (Q1) and third quartile (Q3) for each structure and measurement (CR and nVol). Regions include SNc and N1. Abbreviations: NM, neuromelanin; SNc, substantia nigra; N1, nigrosome-1; nVol, normalised volume; CR, contrast ratio; HC, healthy controls; iPD, idiopathic Parkinson’s disease; GBA-PD, *GBA*-associated Parkinson’s.

**Supplementary Table 3.**
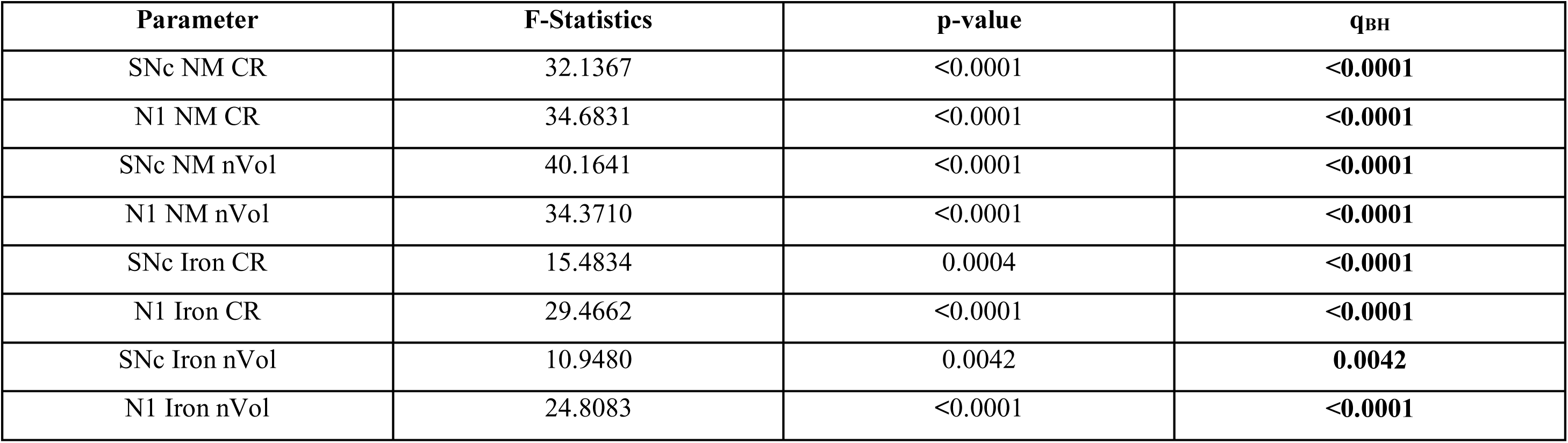
Results of the Kruskal-Wallis global test with Benjamini-Hochberg false discovery rate (FDR) correction applied across all parameters. Values are presented as mean ± SD unless otherwise stated. Statistically significant results after false discovery rate (FDR; Benjamini-Hochberg) correction are shown in bold. Results from the Kruskal-Wallis omnibus tests assessing group-level differences (HC, iPD, and GBA-PD) across NM-and iron-sensitive MRI parameters. Reported statistics include F-values, corresponding p-values, and BH-adjusted qBH values. Significant differences were observed across both contrast ratio (CR) and normalized volume (nVol) metrics, indicating distinct signal alterations in the SNc and N1 among patient groups relative to healthy controls. Abbreviations: NM, neuromelanin; SNc, substantia nigra; N1, nigrosome-1; nVol, normalised volume; CR, contrast ratio; HC, healthy controls; iPD, idiopathic Parkinson’s disease; GBA-PD, *GBA*-associated Parkinson’s.

| Parameter | F-Statistics | p-value | q<sub>BH</sub> |
| --- | --- | --- | --- |
| SNc NM CR | 32.1367 | <0.0001 | **<0.0001** |
| N1 NM CR | 34.6831 | <0.0001 | **<0.0001** |
| SNc NM nVol | 40.1641 | <0.0001 | **<0.0001** |
| N1 NM nVol | 34.3710 | <0.0001 | **<0.0001** |
| SNc Iron CR | 15.4834 | 0.0004 | **<0.0001** |
| N1 Iron CR | 29.4662 | <0.0001 | **<0.0001** |
| SNc Iron nVol | 10.9480 | 0.0042 | **0.0042** |
| N1 Iron nVol | 24.8083 | <0.0001 | **<0.0001** |

**Supplementary Table 4.**
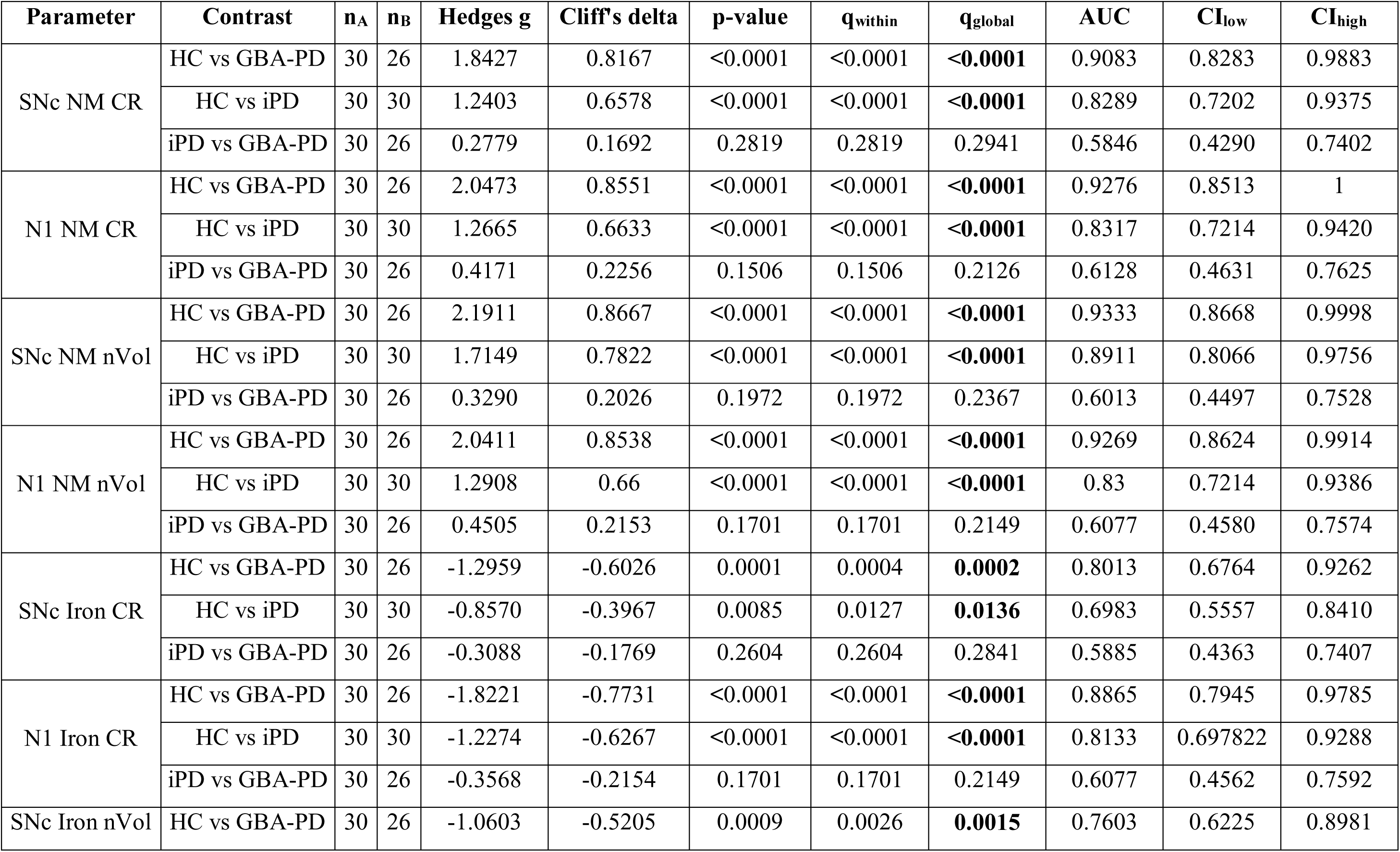
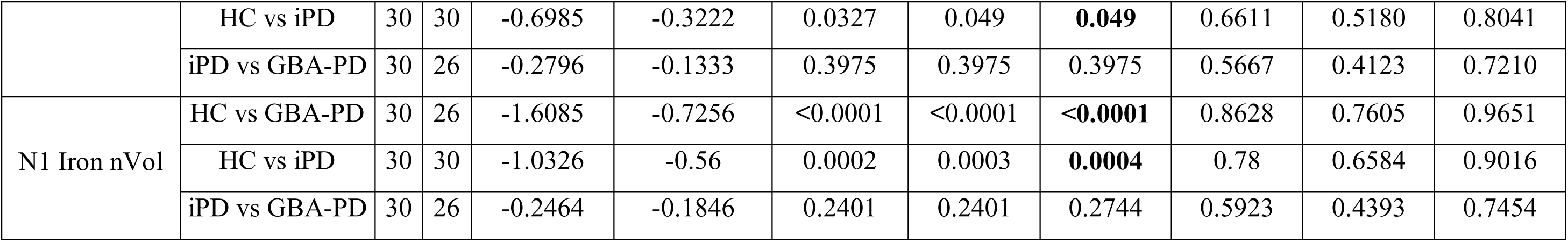
Pairwise comparisons of MRI parameters between groups (BH-corrected) with effect sizes and AUC values with 95%IC. Values are presented as mean ± SD unless otherwise stated. Statistically significant results after false discovery rate (FDR; Benjamini-Hochberg) correction are shown in bold. Pairwise post-hoc comparisons (HC, iPD, and GBA-PD) for NM-and iron-sensitive quantitative MRI parameters across the SNc and N1 subregions, considering both CR and nVol metrics. Reported statistics include Hedges’ g, Cliff’s δ, raw and Benjamini-Hochberg-adjusted p-values, and the area under the receiver operating characteristic curve (AUC) with 95% confidence intervals. Larger effect sizes and AUCs in N1 and SNc NM contrast support the higher discriminative power of NM-sensitive imaging for group differentiation. Abbreviations: NM, neuromelanin; SNc, substantia nigra; N1, nigrosome-1; nVol, normalised volume; CR, contrast ratio; HC, healthy controls; iPD, idiopathic Parkinson’s disease; GBA-PD, *GBA*-associated Parkinson’s.

| Parameter | Contrast | n<sub>A</sub> | n<sub>B</sub> | Hedges g | Cliff's delta | p-value | q<sub>within</sub> | q<sub>global</sub> | AUC | CI<sub>low</sub> | CI<sub>high</sub> |
| --- | --- | --- | --- | --- | --- | --- | --- | --- | --- | --- | --- |
| SNc NM CR | HC vs GBA-PD | 30 | 26 | 1.8427 | 0.8167 | <0.0001 | <0.0001 | **<0.0001** | 0.9083 | 0.8283 | 0.9883 |
|  | HC vs iPD | 30 | 30 | 1.2403 | 0.6578 | <0.0001 | <0.0001 | **<0.0001** | 0.8289 | 0.7202 | 0.9375 |
|  | iPD vs GBA-PD | 30 | 26 | 0.2779 | 0.1692 | 0.2819 | 0.2819 | 0.2941 | 0.5846 | 0.4290 | 0.7402 |
| N1 NM CR | HC vs GBA-PD | 30 | 26 | 2.0473 | 0.8551 | <0.0001 | <0.0001 | **<0.0001** | 0.9276 | 0.8513 | 1 |
|  | HC vs iPD | 30 | 30 | 1.2665 | 0.6633 | <0.0001 | <0.0001 | **<0.0001** | 0.8317 | 0.7214 | 0.9420 |
|  | iPD vs GBA-PD | 30 | 26 | 0.4171 | 0.2256 | 0.1506 | 0.1506 | 0.2126 | 0.6128 | 0.4631 | 0.7625 |
| SNc NM nVol | HC vs GBA-PD | 30 | 26 | 2.1911 | 0.8667 | <0.0001 | <0.0001 | **<0.0001** | 0.9333 | 0.8668 | 0.9998 |
|  | HC vs iPD | 30 | 30 | 1.7149 | 0.7822 | <0.0001 | <0.0001 | **<0.0001** | 0.8911 | 0.8066 | 0.9756 |
|  | iPD vs GBA-PD | 30 | 26 | 0.3290 | 0.2026 | 0.1972 | 0.1972 | 0.2367 | 0.6013 | 0.4497 | 0.7528 |
| N1 NM nVol | HC vs GBA-PD | 30 | 26 | 2.0411 | 0.8538 | <0.0001 | <0.0001 | **<0.0001** | 0.9269 | 0.8624 | 0.9914 |
|  | HC vs iPD | 30 | 30 | 1.2908 | 0.66 | <0.0001 | <0.0001 | **<0.0001** | 0.83 | 0.7214 | 0.9386 |
|  | iPD vs GBA-PD | 30 | 26 | 0.4505 | 0.2153 | 0.1701 | 0.1701 | 0.2149 | 0.6077 | 0.4580 | 0.7574 |
| SNc Iron CR | HC vs GBA-PD | 30 | 26 | -1.2959 | -0.6026 | 0.0001 | 0.0004 | **0.0002** | 0.8013 | 0.6764 | 0.9262 |
|  | HC vs iPD | 30 | 30 | -0.8570 | -0.3967 | 0.0085 | 0.0127 | **0.0136** | 0.6983 | 0.5557 | 0.8410 |
|  | iPD vs GBA-PD | 30 | 26 | -0.3088 | -0.1769 | 0.2604 | 0.2604 | 0.2841 | 0.5885 | 0.4363 | 0.7407 |
| N1 Iron CR | HC vs GBA-PD | 30 | 26 | -1.8221 | -0.7731 | <0.0001 | <0.0001 | **<0.0001** | 0.8865 | 0.7945 | 0.9785 |
|  | HC vs iPD | 30 | 30 | -1.2274 | -0.6267 | <0.0001 | <0.0001 | **<0.0001** | 0.8133 | 0.697822 | 0.9288 |
|  | iPD vs GBA-PD | 30 | 26 | -0.3568 | -0.2154 | 0.1701 | 0.1701 | 0.2149 | 0.6077 | 0.4562 | 0.7592 |
| SNc Iron nVol | HC vs GBA-PD | 30 | 26 | -1.0603 | -0.5205 | 0.0009 | 0.0026 | **0.0015** | 0.7603 | 0.6225 | 0.8981 |
|  | HC vs iPD | 30 | 30 | -0.6985 | -0.3222 | 0.0327 | 0.049 | **0.049** | 0.6611 | 0.5180 | 0.8041 |
|  | iPD vs GBA-PD | 30 | 26 | -0.2796 | -0.1333 | 0.3975 | 0.3975 | 0.3975 | 0.5667 | 0.4123 | 0.7210 |
| N1 Iron nVol | HC vs GBA-PD | 30 | 26 | -1.6085 | -0.7256 | <0.0001 | <0.0001 | **<0.0001** | 0.8628 | 0.7605 | 0.9651 |
|  | HC vs iPD | 30 | 30 | -1.0326 | -0.56 | 0.0002 | 0.0003 | **0.0004** | 0.78 | 0.6584 | 0.9016 |
|  | iPD vs GBA-PD | 30 | 26 | -0.2464 | -0.1846 | 0.2401 | 0.2401 | 0.2744 | 0.5923 | 0.4393 | 0.7454 |

**Supplementary Table 5.** Descriptive statistics and healthy control (HC) reference anchors of asymmetry indices (AI). Values are presented as mean ± SD unless otherwise stated. Statistically significant results after false discovery rate (FDR; Benjamini-Hochberg) correction are shown in bold. Descriptive statistics for asymmetry indices (AI) across groups (HC, iPD, and GBA-PD) for NM-and SWI-based MRI parameters in the SNc and N1 subregions. The table includes raw AI values, p-values, correction status, mean, standard deviation, median, and interquartile range (Q1-Q3). Results indicate moderate asymmetry patterns in NM-and iron-weighted contrasts, particularly within the N1 region, consistent with early dopaminergic lateralization found in PD. Abbreviations: NM, neuromelanin; SNc, substantia nigra; N1, nigrosome-1; nVol, normalised volume; CR, contrast ratio; HC, healthy controls; iPD, idiopathic Parkinson’s disease; GBA-PD, *GBA*-associated Parkinson’s.

| Parameter | Group | n | Raw AI<br>(only<br>HC) | p-value | Correct<br>ion | Mean | SD | Median | Q1 | Q3 |
| --- | --- | --- | --- | --- | --- | --- | --- | --- | --- | --- |
| SNc NM CR<br>AI | HC | 30 | 0.0053 | 0.135 | FALSE | 0.0046 | 0.0154 | 0.0053 | -0.0071 | 0.0129 |
|  | iPD | 29 | - | - | - | 0.0122 | 0.0241 | 0.0162 | -0.0058 | 0.0342 |
|  | GBA-PD | 26 | - | - | - | 0.0027 | 0.0249 | 0.0059 | -0.0119 | 0.0187 |
| N1 NM CR<br>AI | HC | 30 | 0.0621 | **<0.001** | TRUE | -0.0075 | 0.0429 | 0.0001 | -0.0314 | 0.0205 |
|  | iPD | 29 | - | - | - | -0.0516 | 0.0490 | -0.0409 | -0.0731 | -0.0188 |
|  | GBA-PD | 26 | - | - | - | -0.0578 | 0.0739 | -0.0522 | -0.1073 | -0.0260 |
| SNc NM<br>nVol AI | HC | 30 | -0.0031 | 0.556 | FALSE | -0.0092 | 0.0430 | -0.0031 | -0.0238 | 0.0177 |
|  | iPD | 29 | - | - | - | -0.0031 | 0.0560 | -0.0105 | -0.0301 | 0.0381 |
|  | GBA-PD | 26 | - | - | - | -0.0094 | 0.0560 | -0.0034 | -0.0427 | 0.0289 |
| N1 NM nVol<br>AI | HC | 30 | -0.0466 | **0.001** | TRUE | -0.0124 | 0.0991 | 0.0001 | -0.0697 | 0.0338 |
|  | iPD | 29 | - | - | - | 0.0011 | 0.1801 | 0.0171 | -0.1422 | 0.1124 |
|  | GBA-PD | 26 | - | - | - | -0.0089 | 0.1442 | -0.0623 | -0.1193 | 0.1322 |
| SNc Iron CR<br>AI | HC | 30 | 0.0912 | **<0.001** | TRUE | 0.0125 | 0.0855 | 0.0001 | -0.0335 | 0.0574 |
|  | iPD | 29 | - | - | - | 0.0059 | 0.2284 | 0.0282 | -0.1721 | 0.2033 |
|  | GBA-PD | 26 | - | - | - | -0.0501 | 0.2796 | -0.0274 | -0.1889 | 0.1744 |
| N1 Iron CR AI | HC | 30 | 0.53 | **<0.001** | TRUE | -0.0254 | 0.2572 | 0.0001 | -0.1786 | 0.1999 |
|  | iPD | 29 | - | - | - | -0.4504 | 0.5324 | -0.3296 | -0.8294 | -0.0232 |
|  | GBA-PD | 25 | - | - | - | -0.4255 | 0.6098 | -0.2983 | -0.7372 | -0.0514 |
| SNc Iron nVol AI | HC | 30 | -0.009 | 0.477 | FALSE | -0.0183 | 0.0853 | -0.0089 | -0.0538 | 0.0432 |
|  | iPD | 29 | - | - | - | 0.0087 | 0.1041 | 0.0182 | -0.0344 | 0.0537 |
|  | GBA-PD | 26 | - | - | - | 0.0107 | 0.1231 | -0.0001 | -0.0670 | 0.0541 |
| N1 Iron nVol AI | HC | 30 | -0.0646 | 0.981 | FALSE | -0.0257 | 0.4954 | -0.0064 | -0.0944 | 0.2058 |
|  | iPD | 29 | - | - | - | -0.0852 | 0.4542 | -0.0975 | -0.3629 | 0.0883 |
|  | GBA-PD | 26 | - | - | - | -0.0678 | 0.4239 | -0.1500 | -0.3102 | 0.1755 |

**Supplementary Table 6.** Group differences in asymmetry indices (AI). Kruskal-Wallis global test with FDR correction. Results from global Kruskal-Wallis analyses evaluating group-level differences (HC, iPD, and GBA-PD) in asymmetry indices (AI) across NM-and iron-sensitive measures for the SNc and N1 subregions. Statistically significant results after false discovery rate (FDR; Benjamini-Hochberg) correction are shown in bold. Reported statistics include F-values, p-values, degrees of freedom (df = 2), and BH-adjusted qBH. Significant findings were primarily observed in N1 NM and iron contrast ratios, reinforcing the regional specificity of lateralized degeneration patterns. Abbreviations: NM, neuromelanin; SNc, substantia nigra; N1, nigrosome-1; nVol, normalised volume; CR, contrast ratio.

| Parameter | F-Statistics | p-value | q<sub>BH</sub> |
| --- | --- | --- | --- |
| SNc NM CR AI | 3.037 | 0.2190 | 0.5841 |
| N1 NM CR AI | 15.5545 | 0.0004 | **0.0034** |
| SNc NM nVol AI | 0.193 | 0.9080 | 0.9721 |
| N1 NM nVol AI | 0.0565 | 0.9721 | 0.9721 |
| SNc Iron CR AI | 0.5173 | 0.7721 | 0.9721 |
| N1 Iron CR AI | 12.3834 | 0.0020 | **0.0082** |
| SNc Iron nVol AI | 0.8524 | 0.6530 | 0.9721 |
| N1 Iron nVol AI | 1.3741 | 0.5031 | 0.9721 |

**Supplementary Table 7.**
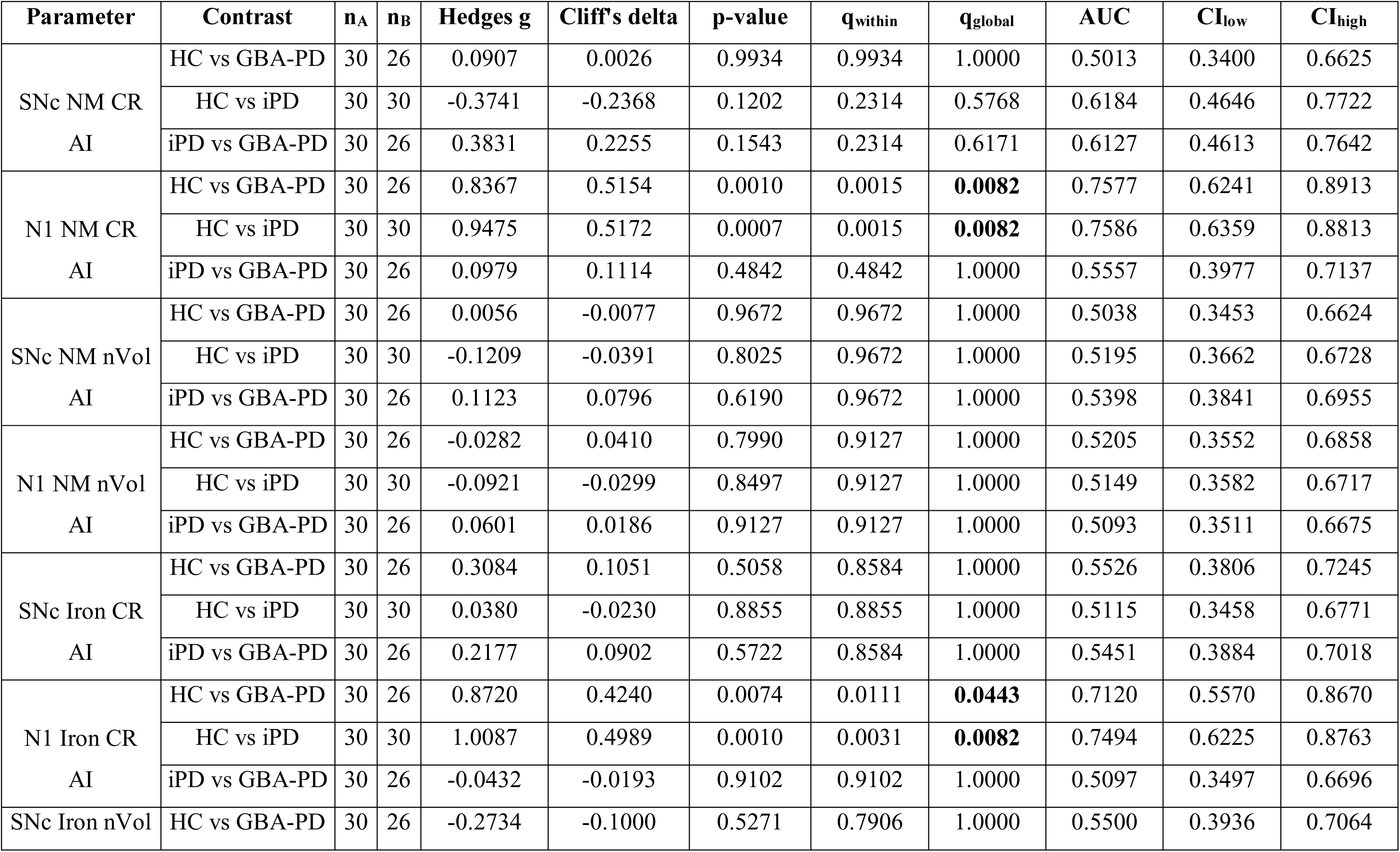
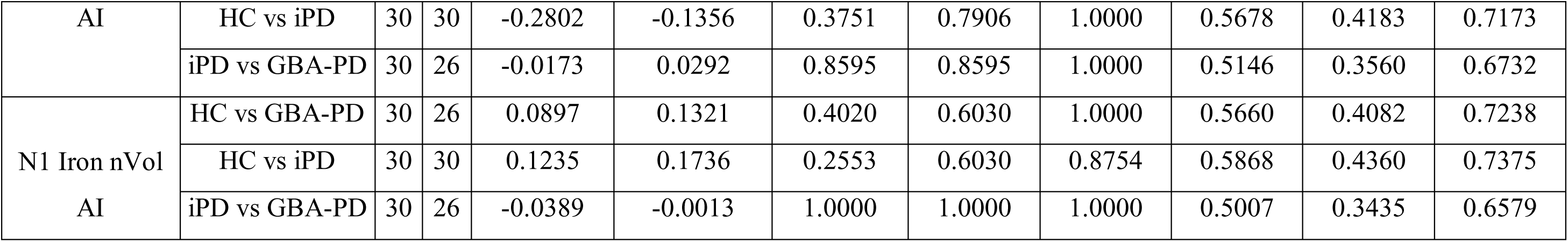
Pairwise comparisons of MRI parameter asymmetry indices (AIs) between groups (BH-corrected) with effect sizes and AUC values with 95% CI. Values are presented as mean ± SD unless otherwise stated. Statistically significant results after false discovery rate (FDR; Benjamini-Hochberg) correction are shown in bold. Pairwise post-hoc comparisons (HC, iPD, and GBA-PD) were performed for asymmetry indices (AI) derived from neuromelanin (NM)-and iron-sensitive (SWI) MRI parameters across the substantia nigra pars compacta (SNc) and nigrosome-1 (N1) subregions, using both contrast ratio (CR) and normalized volume (nVol) metrics. Reported statistics include Hedges’ g, Cliff’s δ, raw and BH-adjusted p-values (pglobal, qglobal), and the area under the receiver operating characteristic curve (AUC) with corresponding 95% confidence intervals (CIlow, CIhigh). Effect-size and AUC profiles indicate subtle but systematic lateralization differences among groups, with the strongest asymmetry shifts observed in N1 NM-related measures, supporting the notion of early laterality for dopaminergic neuronal loss in both idiopathic and GBA-associated Parkinson’s disease. Abbreviations: NM, neuromelanin; SNc, substantia nigra; N1, nigrosome-1; nVol, normalised volume; CR, contrast ratio; HC, healthy controls; iPD, idiopathic Parkinson’s disease; GBA-PD, *GBA*-associated Parkinson’s.

**Supplementary Table 8.** Descriptive statistics of asymmetry indices (AIs) by side of onset in Parkinson’s disease. Values are presented as mean ± SD unless otherwise stated. Statistically significant results after false discovery rate (FDR; Benjamini-Hochberg) correction are shown in bold. Descriptive summary of asymmetry indices (AIs) stratified by clinical side of motor symptom onset (Left vs Right) in idiopathic Parkinson’s disease (iPD) and GBA-associated Parkinson’s disease (GBA-PD). Reported parameters include NM-and iron-sensitive MRI measures from the SNc and N1 subregions, using both CR and nVol metrics. Each entry includes sample size (n), mean, standard deviation (SD), median, and interquartile range (Q1-Q3). Patterns suggest that left-onset and right-onset subgroups exhibit distinct asymmetry distributions, particularly for N1 NM and iron indices, consistent with hemispheric differences in dopaminergic vulnerability in both iPD and GBA-PD cohorts. Abbreviations: NM, neuromelanin; SNc, substantia nigra; N1, nigrosome-1; nVol, normalised volume; CR, contrast ratio; HC, healthy controls; iPD, idiopathic Parkinson’s disease; GBA-PD, *GBA*-associated Parkinson’s.

| Parameter | Group | Onset side | n | Mean | SD | Median | Q1 | Q3 |
| --- | --- | --- | --- | --- | --- | --- | --- | --- |
| SNc NM CR | iPD | Left | 13 | -0.0061 | 0.0224 | -0.0132 | -0.0219 | 0.0058 |
|  |  | Right | 16 | 0.0172 | 0.0250 | 0.0229 | -0.0037 | 0.0391 |
|  | GBA-PD | Left | 16 | -0.0104 | 0.0187 | -0.0097 | -0.0210 | 0.0019 |
|  |  | Right | 10 | -0.0096 | 0.0295 | -0.0129 | -0.0253 | 0.0076 |
| N1 NM CR | iPD | Left | 13 | -0.0252 | 0.0327 | -0.0225 | -0.0564 | -0.0013 |
|  |  | Right | 16 | -0.0016 | 0.0573 | 0.0059 | -0.0369 | 0.0427 |
|  | GBA-PD | Left | 16 | -0.0359 | 0.0641 | -0.0273 | -0.0628 | 0.0039 |
|  |  | Right | 10 | -0.0464 | 0.0608 | -0.0510 | -0.1067 | 0.0063 |
| SNc NM nVol | iPD | Left | 13 | -0.1315 | 0.2077 | -0.1197 | -0.3024 | 0.0474 |
|  |  | Right | 16 | 0.0691 | 0.2469 | 0.1120 | -0.0859 | 0.2481 |
|  | GBA-PD | Left | 16 | -0.1613 | 0.1878 | -0.1661 | -0.3061 | -0.0296 |
|  |  | Right | 10 | -0.1513 | 0.3026 | -0.1800 | -0.3192 | 0.0406 |
| N1 NM nVol | iPD | Left | 13 | -0.4667 | 0.2851 | -0.5064 | -0.6650 | -0.2000 |
|  |  | Right | 16 | -0.2358 | 0.4776 | -0.2721 | -0.5439 | 0.2165 |
|  | GBA-PD | Left | 16 | -0.3711 | 0.3939 | -0.2859 | -0.7340 | -0.0509 |
|  |  | Right | 9 | -0.3706 | 0.6545 | -0.7189 | -0.7424 | -0.1629 |
| SNc Iron CR | iPD | Left | 13 | 0.0407 | 0.0448 | 0.0260 | 0.0105 | 0.0826 |
|  |  | Right | 16 | 0.0275 | 0.0451 | 0.0282 | -0.0055 | 0.0551 |
|  | GBA-PD | Left | 16 | 0.0092 | 0.0621 | -0.0066 | -0.0244 | 0.0506 |
|  |  | Right | 10 | -0.0099 | 0.0477 | -0.0151 | -0.0211 | 0.0289 |
| N1 Iron CR | iPD | Left | 13 | 0.1901 | 0.1112 | 0.1932 | 0.1714 | 0.2657 |
|  |  | Right | 16 | 0.0719 | 0.1335 | 0.0258 | -0.0271 | 0.1681 |
|  | GBA-PD | Left | 16 | 0.0566 | 0.1333 | 0.0890 | -0.0719 | 0.1684 |
|  |  | Right | 10 | -0.0539 | 0.1678 | -0.1089 | -0.1523 | 0.0929 |
| SNc Iron nVol | iPD | Left | 13 | 0.0512 | 0.0892 | 0.0344 | -0.0157 | 0.0839 |
|  |  | Right | 16 | 0.0573 | 0.0909 | 0.0480 | 0.0176 | 0.0815 |
|  | GBA-PD | Left | 16 | -0.0212 | 0.1396 | 0.0263 | -0.0810 | 0.0727 |
|  |  | Right | 10 | -0.0061 | 0.0953 | 0.0342 | -0.0381 | 0.0540 |
| N1 Iron nVol | iPD | Left | 13 | 0.2925 | 0.4033 | 0.3629 | 0.0807 | 0.4024 |
|  |  | Right | 16 | 0.0832 | 0.4330 | -0.0249 | -0.1448 | 0.2847 |
|  | GBA-PD | Left | 16 | -0.0330 | 0.4747 | -0.0544 | -0.2264 | 0.1753 |
|  |  | Right | 10 | -0.2291 | 0.2767 | -0.1957 | -0.3880 | -0.1453 |

**Supplementary Table 9.**
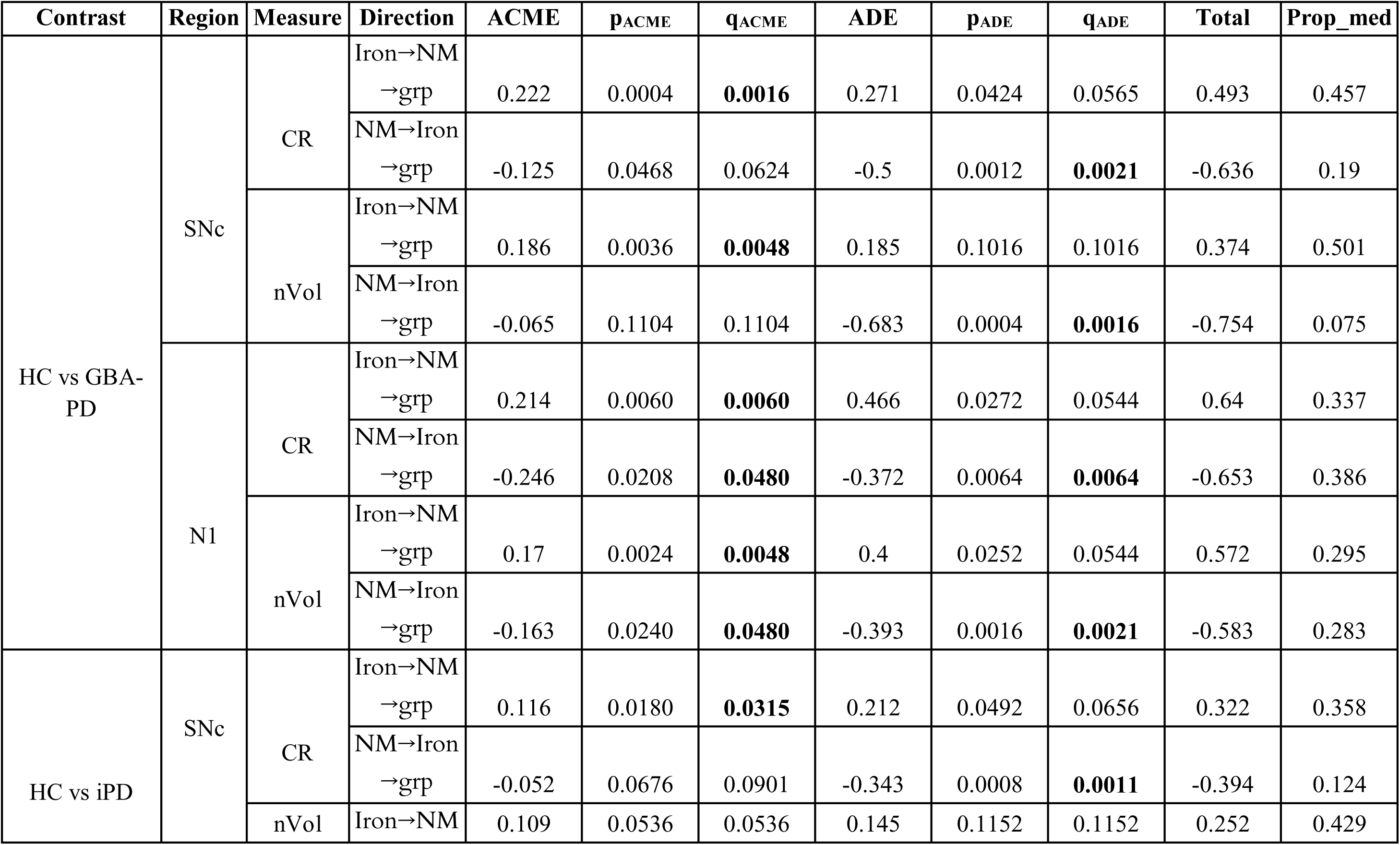
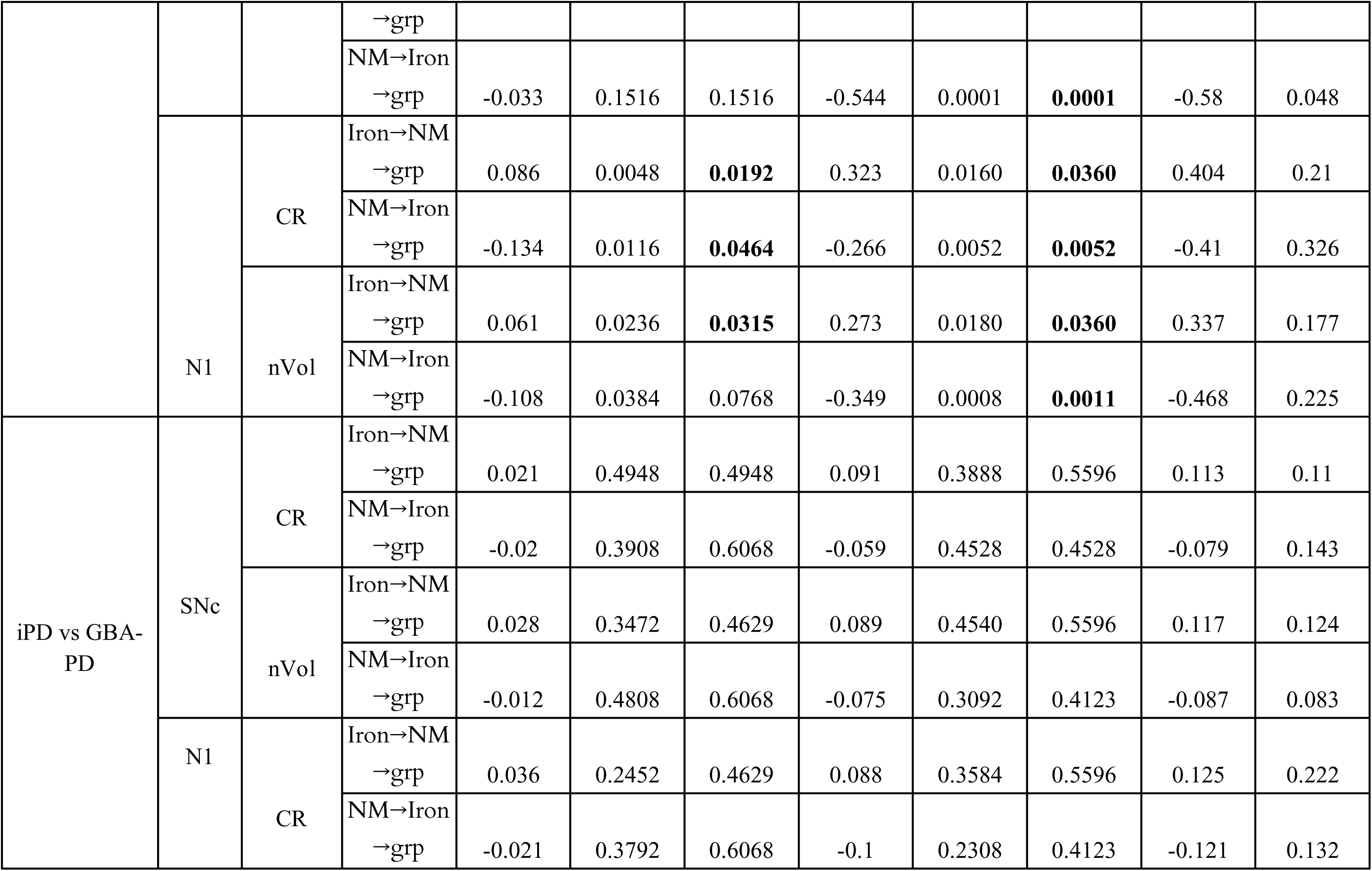
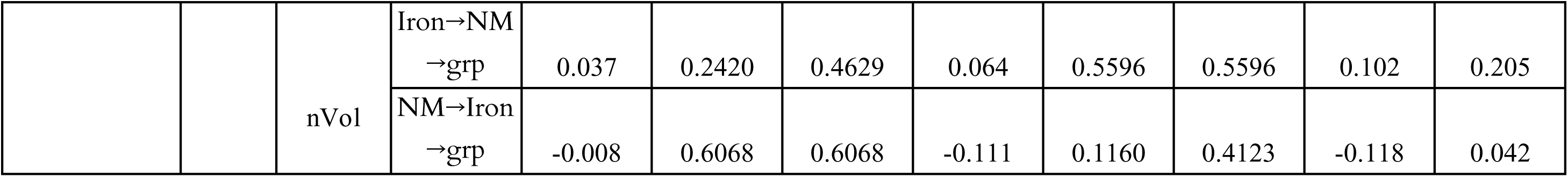
Bidirectional mediation analyses of NM-iron coupling across regions and groups. Values are presented as unstandardized coefficients (ACME, ADE, and total effects), with associated p-values and Benjamini-Hochberg-adjusted q-values. Statistically significant results after FDR correction are shown in bold. Mediation analyses were conducted to examine the directional interplay between NM and iron-sensitive MRI metrics-tested bidirectionally (Iron→NM→Group and NM→Iron→Group)-in the SNc and N1 subregions, using both CR and nVol measures. Each comparison (HC vs GBA-PD, HC vs iPD, iPD vs GBA-PD) reports the average causal mediation effect (ACME), average direct effect (ADE), total effect, and the proportion mediated (prop_med), together with p-and q-values for each component. Significant indirect effects (ACME) were predominantly observed in NM-mediated pathways (NM→Iron→Group) for the HC vs GBA-PD and HC vs iPD contrasts-particularly within the N1 and SNc subregions-indicating that group differences in iron accumulation may be partially explained by variations in NM signal. Conversely, direct effects (ADE) remained stronger in Iron→NM→Group models, suggesting that iron-sensitive changes independently contribute to group differentiation. These patterns support a bidirectional but asymmetrical coupling between NM and iron processes, with NM-to-iron mediation emerging as a dominant pathway in moderated iPD and genetic PD phenotypes. Abbreviations: NM, neuromelanin; SNc, substantia nigra; N1, nigrosome-1; nVol, normalised volume; CR, contrast ratio; HC, healthy controls; iPD, idiopathic Parkinson’s disease; GBA-PD, *GBA*-associated Parkinson’s.

**Supplementary Table 10.**
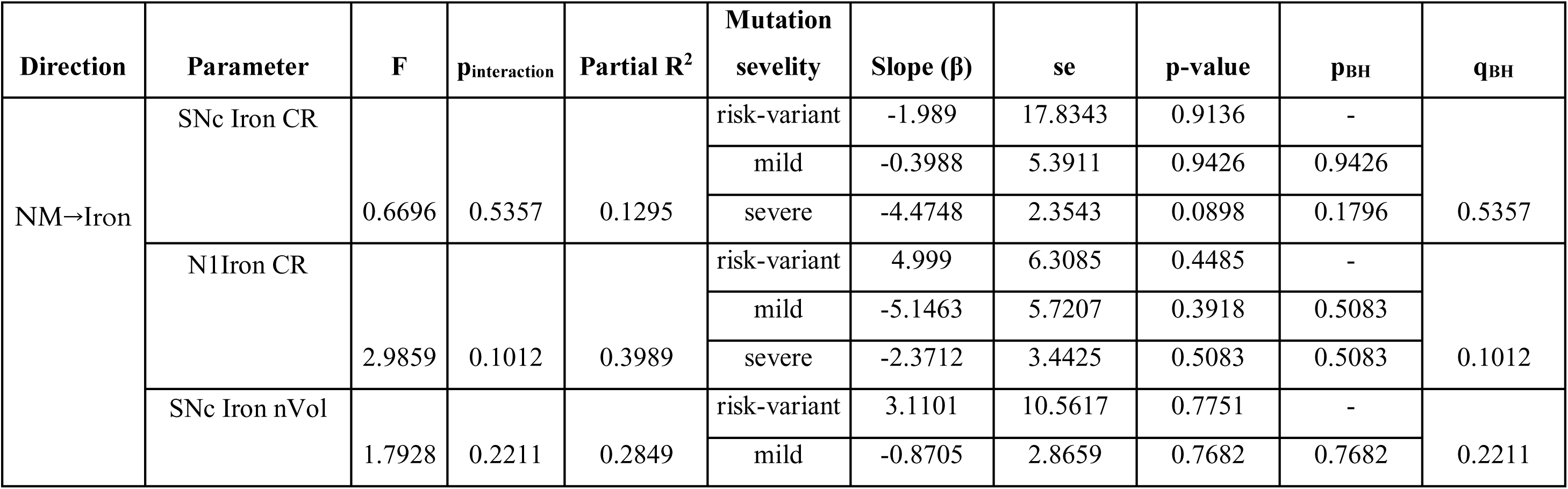
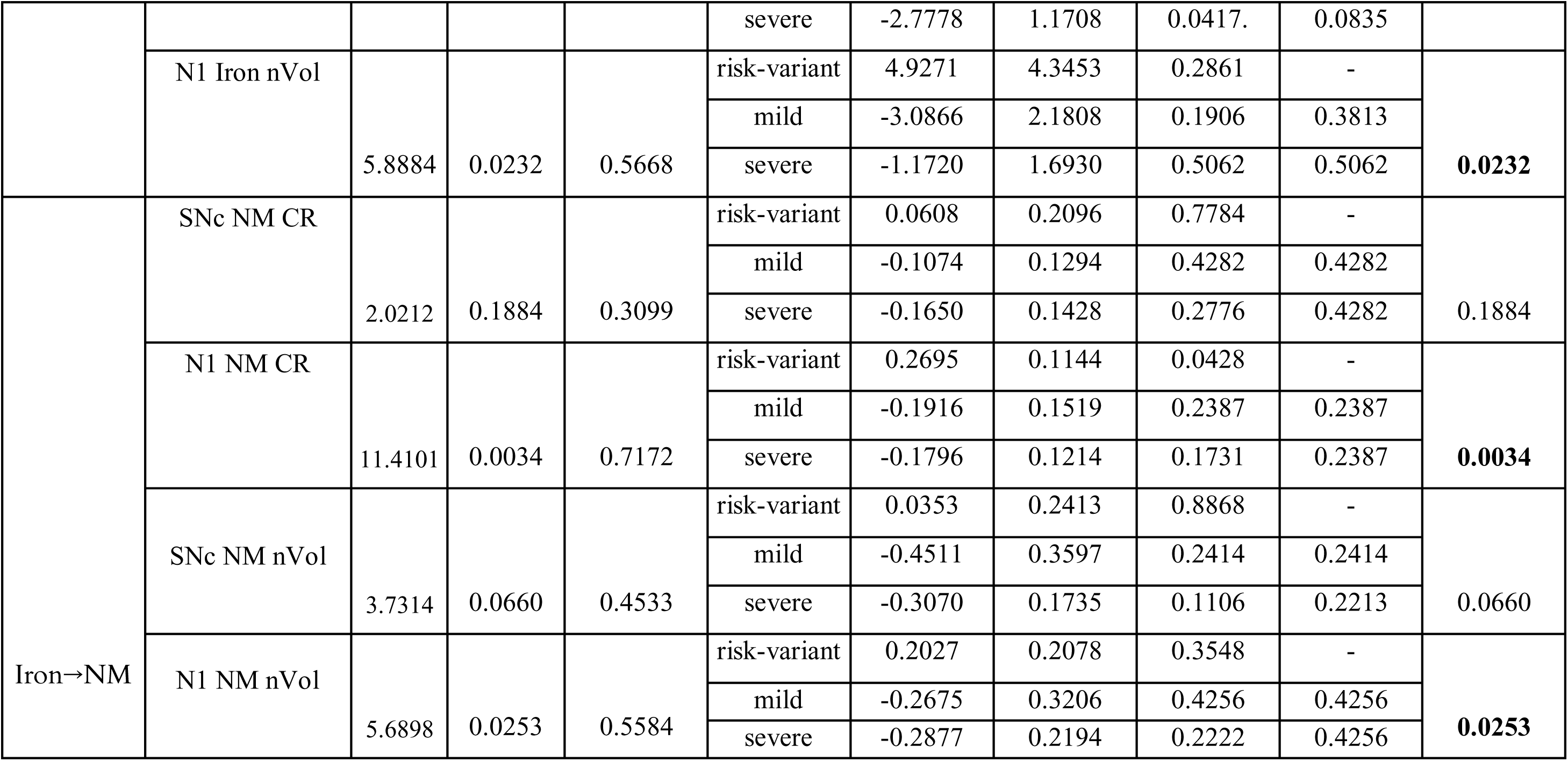
Moderation analyses by GBA mutation severity in GBA-PD. Values are presented as unstandardized regression coefficients (β) with standard error (SE), p-values, and Benjamini-Hochberg-adjusted q-values. Statistically significant results after FDR correction are shown in bold. Moderation analyses were performed exclusively within the GBA-PD cohort to test whether mutation severity (risk-variant, mild, severe) modulated the bidirectional coupling between NM and iron-sensitive MRI parameters in the SNc and N1, using both CR and nVol. The models included covariates (age, sex, onset side, UPDRS, Hoehn & Yahr stage, MMSE, education, disease duration, and LEDD). Reported values comprise F-statistics, interaction p-values (pinteraction), partial R², and simple slopes for each severity category. Significant moderation emerged primarily in N1 for both NM and iron parameters, with stronger NM↔iron coupling in risk-variant carriers and attenuated or reversed associations in mild and severe mutation groups. These findings suggest that mutation severity shapes the balance between NM loss and iron accumulation, potentially reflecting distinct molecular trajectories of nigral degeneration in GBA-associated Parkinson’s disease. Abbreviations: NM, neuromelanin; SNc, substantia nigra; N1, nigrosome-1; nVol, normalised volume; CR, contrast ratio.

**Supplementary Table 11.**
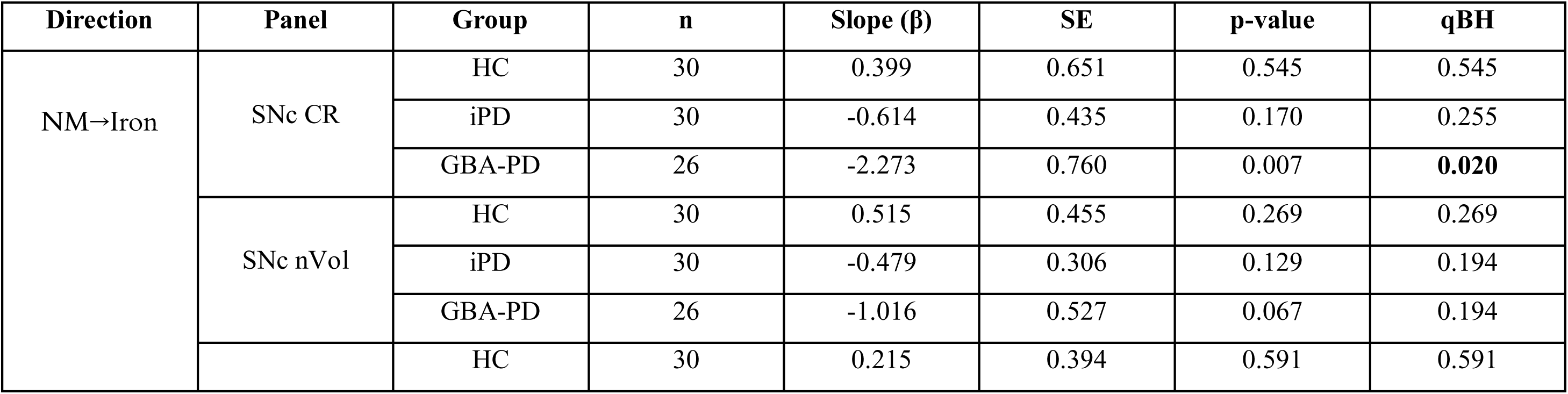
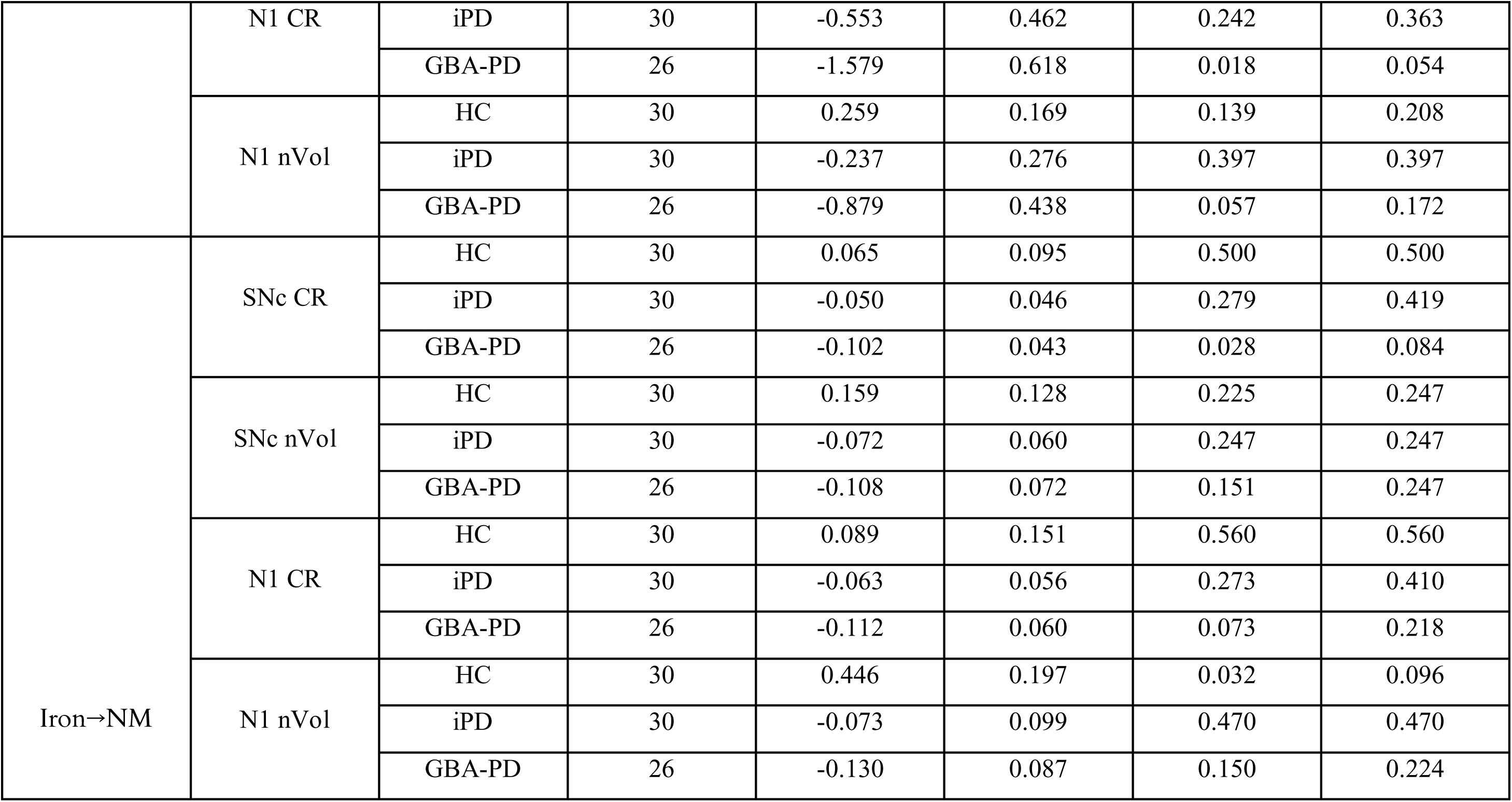
Slopes of NM-iron coupling across groups and regions. Each slope (β) represents the strength of the linear association between NM and iron-sensitive MRI indices within each group-healthy controls (HC), idiopathic Parkinson’s disease (iPD), and monogenic GBA-associated PD (GBA-PD)-adjusted for age and sex using heteroskedasticity-robust (HC2) standard errors. Positive β values indicate that higher NM signal is associated with higher iron estimates, whereas negative β values indicate an inverse relationship (reduced NM accompanied by elevated iron). Significant effects (in bold) emerged exclusively in the GBA-PD group, showing a strong negative NM→Iron coupling in the SNc (CR, β =-2.27, p = 0.007, q = 0.02) and a trend in N1 (CR, β =-1.58, p = 0.018, q = 0.054), consistent with enhanced NM-iron dysregulation. Reverse associations (Iron→NM) were weaker, with only a modest negative slope in GBA-PD (SNc CR, p = 0.028). All p-values were two-tailed; qBH denotes the Benjamini-Hochberg false discovery rate. Abbreviations: NM, neuromelanin; SNc, substantia nigra; N1, nigrosome-1; nVol, normalised volume; CR, contrast ratio; HC, healthy controls; iPD, idiopathic Parkinson’s disease; GBA-PD, *GBA*-associated Parkinson’s.

**Supplementary Table 12.**
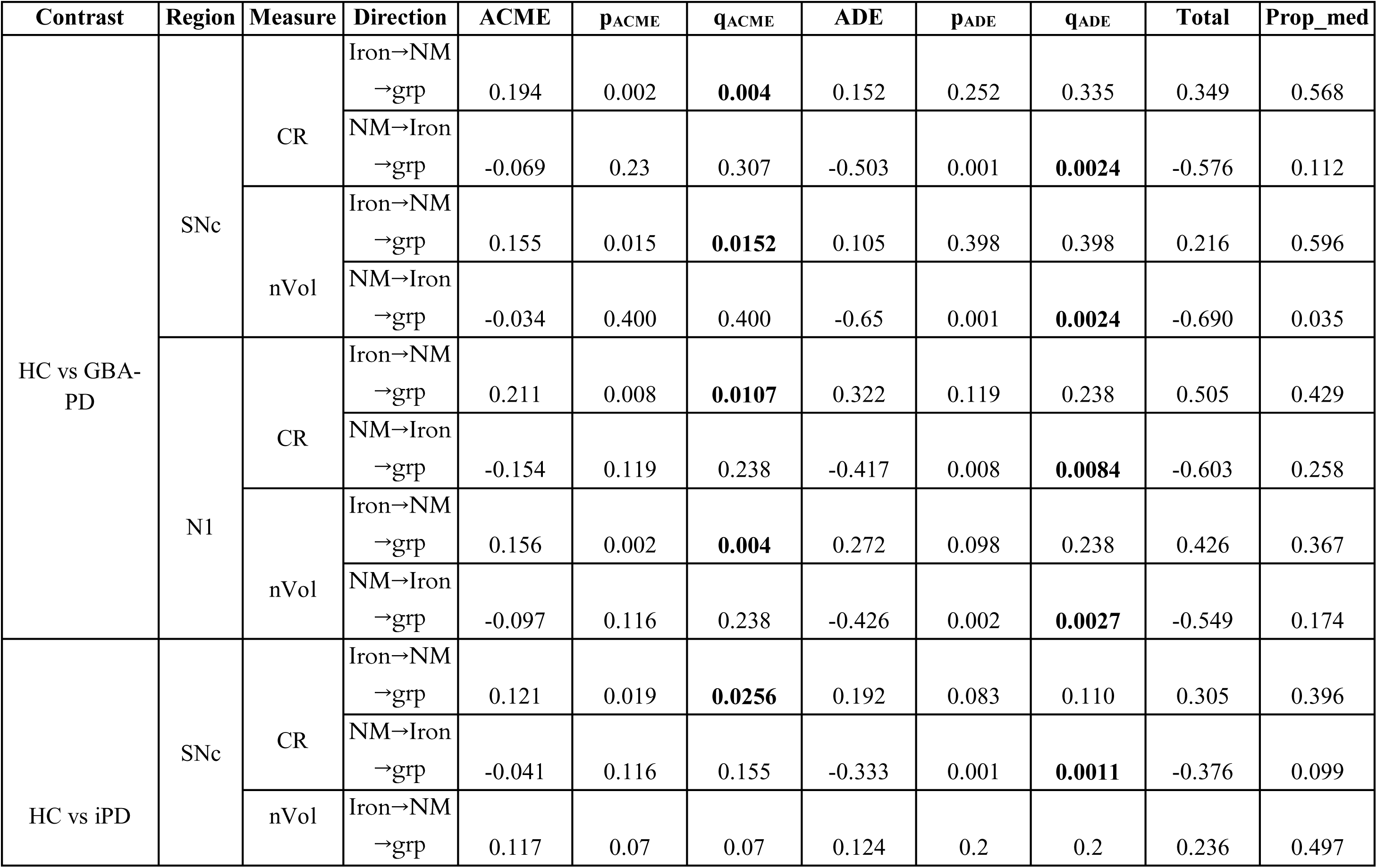
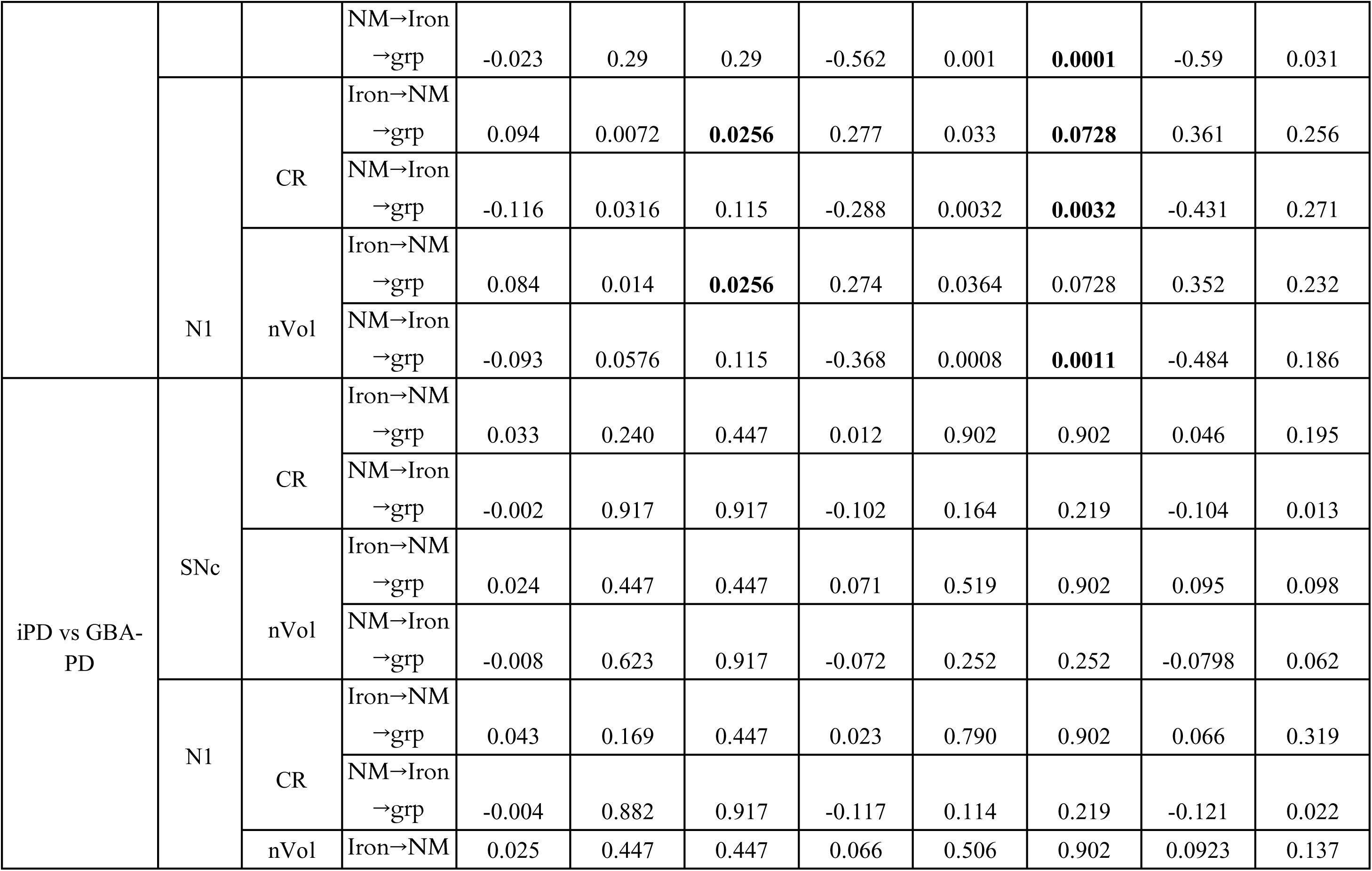
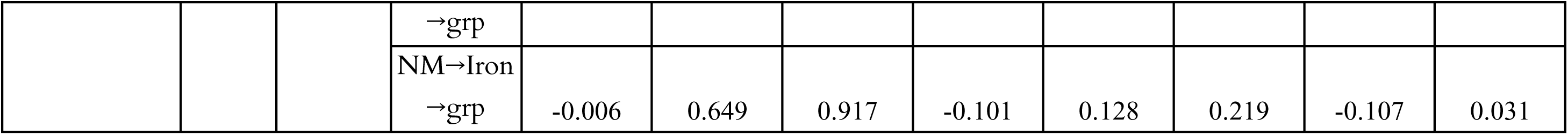
Sensitivity analyses for mediation models. Values represent the robustness of mediation pathways under parameter uncertainty using quasi-Bayesian estimation with 5,000 simulations. Reported coefficients include the average causal mediation effect (ACME), average direct effect (ADE), total effect, and proportion mediated (prop_med), and adjusted q-values (Benjamini-Hochberg correction). Sensitivity analyses were performed for all significant and trend-level mediation models described in Supplementary Table 9, across NM and iron-sensitive MRI measures (CR, nVol) in the SNc and N1. The directionality of mediation was tested bidirectionally (Iron → NM → Group and NM → Iron → Group). Overall, results confirmed the stability of NM-mediated effects observed in the main models (HC vs GBA-PD > HC vs iPD), particularly within N1, where indirect effects (ACME) remained significant after bootstrapping and multiple-comparison correction. The relative proportion of mediated variance indicates that a substantial fraction of group differences in iron signal is explained by NM-linked pathways. No significant deviations or model instabilities were detected under sensitivity testing, supporting the robustness of the NM-iron bidirectional mediation framework. Abbreviations: NM, neuromelanin; SNc, substantia nigra; N1, nigrosome-1; nVol, normalised volume; CR, contrast ratio; HC, healthy controls; iPD, idiopathic Parkinson’s disease; GBA-PD, *GBA*-associated Parkinson’s.

